# *CREBRF* rs373863828 minimally regulates gene expression in subcutaneous adipose tissue of Samoan adults

**DOI:** 10.64898/2026.09.21.26363571

**Authors:** Samantha L. Manna, Anna C. Rivara, Angela T. Prescott, Corina S. Penaia, Muagututia Sefuiva Reupena, Take Naseri, Satupaitea Viali, Melania Selu, Gloria Siufaga, Kima Faasalele-Savusa, Folla Unasa, Vaimoana Lupematisila, Alysa Pomer, Elizabeth C. Lawrence, Jenna C. Carlson, Emily M. Russell, Mohanraj Krishnan, Katie L. Whytock, Lacey W. Heinsberg, Erin E. Kershaw, Ryan L. Minster, Daniel E. Weeks, Stephen T. McGarvey, Nicola L. Hawley, Zsolt Urban

**Affiliations:** Center for Craniofacial and Dental Genetics, Department of Oral and Craniofacial Sciences, School of Dental Medicine, University of Pittsburgh, Pittsburgh, PA, USA; Department of Human Genetics, School of Public Health, University of Pittsburgh, Pittsburgh, PA, USA; Department of Social and Behavioral Sciences, O’Donnell School of Public Health, University of Texas Southwestern Medical Center, Dallas, TX, USA; Department of Plastic and Reconstructive Surgery, University of Pittsburgh Medical Center, Pittsburgh, PA, USA; Department of Health Policy and Management, Fielding School of Public Health, University of California, Los Angeles, Los Angeles, CA, USA; Lutia I Puava Ae Mapu I Fagalele, Apia, Samoa; Ministry of Health, Apia, Samoa; Oceania University of Medicine, Samoa; Department of Chronic Disease Epidemiology, Yale School of Public Health, New Haven, CT, USA; Obesity, Lifestyle and Genetic Adaptations Study Group, Apia, Samoa; Center for Surgery and Public Health, Brigham and Women’s Hospital, Boston, MA, USA; Department of Biostatistics and Health Data Science, School of Public Health, University of Pittsburgh, Pittsburgh, PA, USA; Department of Biobehavioral Health, Pennsylvania State University, State College, PA, USA; Department of Epidemiology, University of North Carolina at Chapel Hill, Chapel Hill, NC, USA; Translational Research Institute, AdventHealth, Orlando, Florida, USA; Department of Health Promotion and Development, School of Nursing, University of Pittsburgh, Pittsburgh, PA, USA; Division of Endocrinology and Metabolism, Department of Medicine, School of Medicine, University of Pittsburgh, Pittsburgh, PA, USA; Center for Global Public Health, Department of Epidemiology, Brown University School of Public Health, Providence, RI, USA; Department of Anthropology, Brown University, Providence, RI, US

## Abstract

A population-specific missense variant, rs373863828 (G>A, p.Arg457Gln), has been identified in Pacific Islanders and is associated with increased body mass index and lower odds of type 2 diabetes. However, the mechanism underlying these associations is unknown. The variant is located in exon 5 of *CREBRF*, a gene which is rapidly upregulated in response to nutrient deprivation. This study aimed to assess the effect of rs373863828 on gene expression in adipose tissue from Samoan adults from the *Soifua Manuia* (“Good Health”) study. Needle subcutaneous adipose (upper buttock) biopsies were obtained in Apia, Samoa, from fasting Samoans without diabetes. RNA isolated from adipose tissue (n=91, 65.9% female) was subjected to bulk paired-end mRNA-sequencing (2×150 base pair). Differential gene expression among the three rs373863828 genotypes (GG, AG, AA) was assessed using a likelihood-ratio test implemented in DESeq2, and overrepresentation analysis for significant differentially expressed genes (DEGs) (*p_adj_* < 0.1) was performed with clusterProfiler and oRG.DB. Bulk expression was deconvoluted to determine the cell type composition of the adipose tissue. Twenty-two DEGs were associated with rs373863828 genotype including *CBX7* (*p_adj_*=0.026) and *SELENOM* (*p_adj_*= 0.004), both of which have roles in adipogenesis and metabolism. A single expression profile comprising 13 of the 22 DEGs corresponding to *CREBRF* genotype was identified, and overrepresentation analysis supported this finding, with terms such as “keratinocyte differentiation” and “skin development” enriched among significant DEGs. However, PERMANOVA of adipose cell type proportions showed no difference (*p* > 0.05) among the three genotypes. rs373863828 genotype was associated with expression of 22 genes in whole subcutaneous adipose tissue of this sample of fasting Samoan adults without diabetes. Future work should assess cell- and tissue-specific rs373863828 genotype effects using a combination of single-cell approaches and human cell models with perturbations associated with nutritional stress.

**AUTHOR SUMMARY:** A genetic variant (change in DNA, G >A) is associated with higher body mass index and lower risk for type 2 diabetes. This variant, called rs373863828, is only present in people of Pacific Islander ancestry, who are underrepresented in genetic and genomic studies, and is in the *CREBRF* gene, which is not well-understood. Studies in mice, flies, and cells from humans and mice suggest *CREBRF* is important for cellular growth and stress response, and that rs373863828 increases these functions. In this study, we tested the hypothesis that *CREBRF* affects these cellular processes by controlling gene expression—how genes are “turned on or off” –in fat tissue from Samoan adults as part of the *Soifua Manuia* (“Good Health”) study. We found 22 genes that were affected by how many G to A changes (0, 1, or 2) an individual has at this position in *CREBRF*. We also found that the variant did not affect the proportion of the cell types that make up adipose tissue. This work helps us to better understand the function of the *CREBRF* gene in human fat and gives some clues about how the variant affects BMI and type 2 diabetes risk in healthy Samoan adults.

## INTRODUCTION

A missense variant (rs373863828 G>A, p.Arg457Gln) in the gene encoding putative transcriptional regulator CREB3 Regulatory Factor, *CREBRF,* is associated with higher mean body mass index (BMI), taller stature, greater lean mass, and lower fat mass in individuals of Pacific Islander ancestry, but is largely absent from other global populations (1–9). Overweight/obesity (OW/OB) is a well-established risk factor for type 2 diabetes (T2D), and rates of both conditions have been increasing steadily in Samoa since at least 1978 (10–12). These increases are largely attributable to the nutrition transition and, while the effect size of rs373863828 for mean BMI is one of the largest seen for a common variant associated with this phenotype (∼1.4 kg/m^2^ per copy of alternate allele), the variant alone does not account for the epidemic rates of OW/OB in this population [9–11]. Furthermore, the rs373863828 minor allele (A) is associated with *lower* odds of T2D and fasting glucose despite its association with higher mean BMI (1,5,7,13).

While associations of OW/OB and T2D with this variant are clear and robust, the mechanism(s) by which it produces them remains less so. Characterization of *CREBRF* function has been minimal, with roles in autophagy, glucocorticoid signaling, maternal behavior, embryo implantation, and nutritional stress response described (1,14–22). Evidence in *Drosophila* indicates orthologous *REPTOR* is not only responsive to nutritional environment, but also plays a pivotal role in metabolic flexibility and energy substrate choice in muscle (15,23).

Adipose tissue is critical for energy homeostasis, with direct implications for BMI and T2D risk, and the association of rs373863828 with these same phenotypes makes it a strong candidate in which to investigate how gene expression is affected by rs373863828 (24). Thus, we aimed to identify associations of rs373863828 with expression of downstream gene targets in the adipose tissue of Samoan adults without diabetes.

## METHODS

### Subjects and Biopsy Procedure

Adipose biopsies were collected in January and February 2019 in Apia, Samoa, from participants in the *Soifua Manuia* (“Good Health”) Study as previously described (25,26). The *Soifua Manuia* study captured additional metabolic, behavioral, and anthropometric phenotypes with the aim of better characterizing the role of rs373863828 in metabolic outcomes. Briefly, 118 participants who, at the time of enrollment in the primary *Soifua Manuia* study, 1) reported no history of T2D, 2) had HbA1c < 6.5%, and 3) consented to the adipose biopsy were recruited for the procedure. Recruitment for the biopsy study targeted individuals without a history of cancer or cardiovascular disease, with equal distributions of male and female participants across all three rs373863828 genotypes (GG, AG, AA). Individuals who reported pregnancy, lactation, or birth within the last 6 months were excluded. Given the time between recruitment into the primary *Soifua Manuia* study and the biopsy procedure, participants in the biopsy study were re-screened for exclusion criteria, including HbA1c% (A1cNow+ kits, PTS Diagnostics) or fasting glucose levels (Bayer Contour Next point-of-care devices) the day before the biopsy. For participants who 1) reported eating within 2 hours of re-screening, 2) had HbA1c% ≥ 6.5, or 3) had fasting glucose between 126 mg/dl and 140 mg/dl at re-screening, fasting glucose was measured in triplicate the next morning. Participants were eligible for the procedure if the average was ≤ 126 mg/dl. All participants were verbally re-consented by research assistants in the Samoan language at the time of the procedure. Approximately 100 mg of subcutaneous white adipose tissue (sWAT) was collected from the upper buttock via needle and syringe after the anticipated biopsy site was initially numbed with an ice pack, cleaned with single-use sterile ChloraPrep^TM^ (Becton, Dickinson and Company) and additionally numbed with topical Gebauer’s ethyl chloride spray. All biopsies were performed in a medical setting by the same physician (A. Prescott), and the tissue was stored in 0.5-1 ml RNA*later* (Ambion Inc., Waltham, MA, USA) per the manufacturer’s protocol. A total of 118 individuals were biopsied, including seven individuals who had a re-screen HbA1c ≥ 6.5% due to a miscommunication of exclusion criteria. These participants were sequenced but not included in differential gene expression analyses (**Fig 1**). Institutional review board (IRB) review and approval were granted by Yale University (IRB #1604017547), University of Pittsburgh (IRB #PRO16040077), and the Health Research Committee of Samoa.

**Figure 1.**
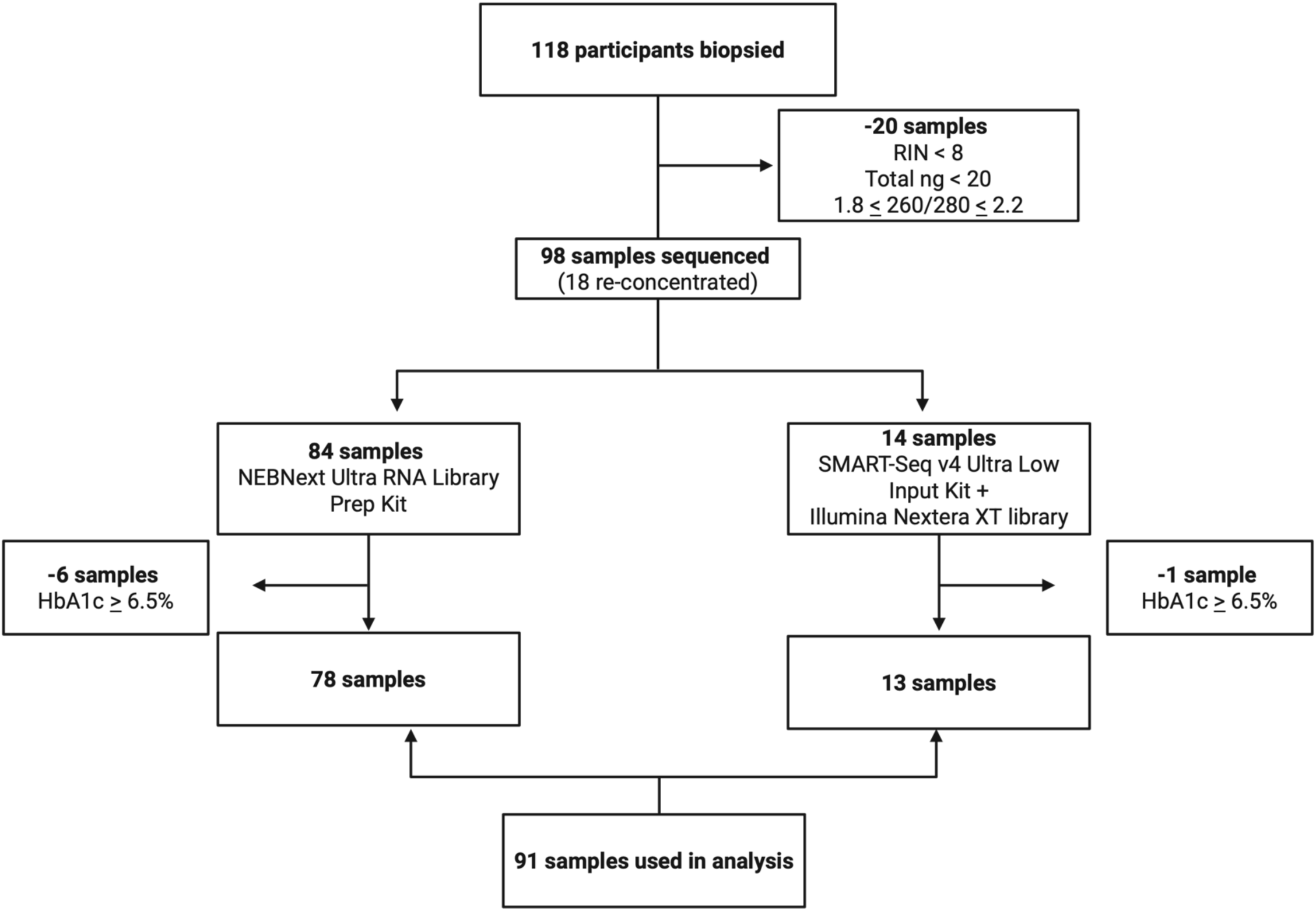
Consort diagram of participants biopsied, quality control, and final analytical sample. RIN: RNA integrity number. Created in BioRender. Created in BioRender. Manna, S. (2027) https://BioRender.com/7huggre

### RNA Isolation and Sample Quality Control

Total RNA was isolated as described (25) using RNeasy Lipid Tissue Mini Kit with on-column DNase I treatment (QIAGEN, Germantown, Maryland, USA) and quantified with a Qubit 3.0 Fluorometer (Invitrogen, Waltham, MA, USA). Protein contamination (A_260/280_) was measured using a Nanodrop spectrophotometer (ThermoFisher, Waltham, MA, USA). The University of Pittsburgh Genomics Research Core assessed RNA integrity number (RIN) and DV200 using TapeStation 4200 (Agilent, Santa Clara, CA, USA). Eighteen samples with concentrations < 20 ng/ul were concentrated with a DNA110 Speedvac Concentrator (Savant) on the low setting for 30 minutes.

### Sequencing

mRNA sequencing was performed by Genewiz (South Plainfield, NJ, USA). Based on quality control at the University of Pittsburgh and by Genewiz, 98 of 118 (83%) samples had high-quality (RIN ≥ 8, 260/280: 1.8-2.2) RNA for sequencing, and 84 of those samples had sufficient ( ≥ 500 ng, ≥ 50 ng/ul) for standard library preparation. Briefly, NEBNext Ultra RNA Library Prep Kit for Illumina (NEB, Ipswich, MA, USA) was used to generate unstranded RNA sequencing libraries for 2×150 base pair (bp) paired-end sequencing with Illumina HiSeq. For samples with fewer than 500 ng of RNA or concentrations less than 50 ng/ul (n=14), SMART-Seq v4 Ultra Low Input Kit for Sequencing was used for full-length cDNA synthesis and amplification (Clontech, Mountain View, CA), and Illumina Nextera XT DNA Library Prep Kit was used for sequencing library preparation. Samples were sequenced to a mean depth of 25.5 million reads.

Sequencing adapters were trimmed with Trimmomatic (27) (v.0.39) using a 4-base sliding window with read quality of ≥ 15 and a minimum length of 36 bp. Quality for both trimmed and untrimmed reads was assessed with FastQC (28) (v.0.11.8). Following adapter trimming, paired reads were aligned to GRCh38 reference genome with ENSEMBL annotation and quantified with STAR aligner (29) (v.2.7.0e, --quantMode GeneCounts). Alignment files (BAM) were indexed with SAMtools (v.1.9) (30), and quality control for read alignment was performed using RSeQC (31) (v.3.0.0). Uniquely mapped reads averaged 92.7% across all samples used in this analysis.

### Differential Gene Expression (DGE) Analysis

The final sample used for the analyses described was 91 (65.9% female; **Table 1**, **Fig 1**; 81% OW/OB (BMI > 30 kg/m^2^), **Supp Fig 1**). Exploration of variance stabilized expression principal components showed the main drivers of variation at the sample or participant level were library preparation (PC1, 56%) and sex (PC2, 12%); thus we used both as covariates in our model (∼sex + library + rs373863828 genotype) (**Fig 2**). A likelihood ratio test (LRT) implemented in DESeq2 (32) (v. 1.46.0) with rs373863828 genotype as the main effect was used to identify differentially expressed genes (DEGs) across the 3 genotypes (GG, AG, AA). Only genes with a minimum of 10 reads in at least 10% of the samples were included in the LRT. Results were annotated using bioMart (33,34) (v. 2.62.1). A gene was considered to be differentially expressed if the Benjamini–Hochberg adjusted p value < 0.10, and DEGreport (v.1.42.0) (35) was used to identify shared expression profiles among DEGs. Overrepresentation analysis of significant DEGs was performed using clusterProfiler (36,37) (v. 4.14.6) and org.Hs.eg.db (v.3.20.0) (38), and expression of these genes in skin, adipose tissue, and cultured fibroblasts was queried in the Adult Genotype Tissue Expression (GTEx) portal [accessed 13 August 2024]. All analyses were performed in R/RStudio Server (39) (v. 4.4.1/2024).

**Table 1:** Subject demographics by rs373863828 genotype.

|  | GG | AG | AA |
| --- | --- | --- | --- |
| N (% female) | 33 (75.8%) | 34 (58.8%) | 24 (62.5%) |
| Age (years) | 50.16 (33.87 - 67.51) | 51.81 (35.98 - 70.25) | 52.31 (39.42 - 71.36) |
| BMI (kg/m^2) | 35.36 (23.24 - 46.36) | 37.98 (23.66 - 63.26) | 36.76 (26.56 - 57.83) |
| HbA1c (%) | 5.9 (5.3 - 6.4) | 5.83 (5.3 - 6.3) | 5.85 (5.1 - 6.4) |

\* Data are mean (range) unless otherwise indicated.

**Figure 2.**
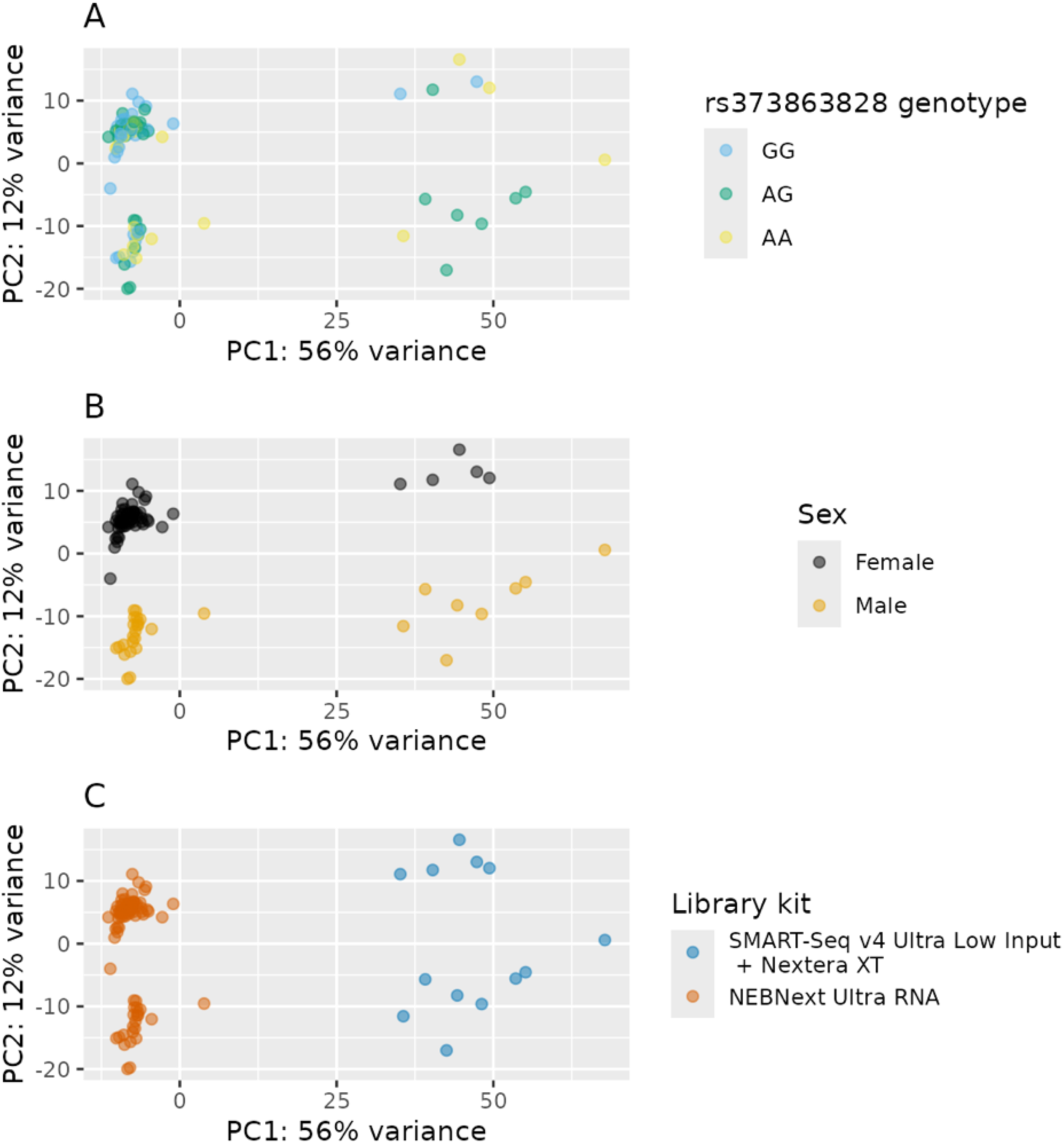
Principal components analysis of variance stabilized expression. indicate rs373863828 genotype is not a major driver of variance in gene expression **(A)**, but participant sex **(B)** and sequencing library kit **(C)** are.

### Deconvolution

Given the heterogeneity of adipose tissue, we performed deconvolution of expression data as described by Whytock, *et al,* to determine if cell type proportions differed by rs373863828 genotype (40,41). Briefly, DESeq2 normalized read counts were deconvoluted using a full-length single nuclei transcriptomic reference matrix derived from 20 subcutaneous adipose tissue samples from an external, non-Samoan dataset. Cell types in the matrix included adipose tissue cell types: stem cells, macrophages, mast cells, pre-adipocytes, vascular cells, and two types of adipocytes, Adip_1 (adipocytes enriched for oxidation and immune response genes) and Adip_2 (adipocytes enriched for insulin signaling and lipid metabolism). Following deconvolution, we compared cell type proportions by rs373863828 genotype using PERmutational Multivariate ANalysis of VAriance (PERMANOVA) as implemented by the adonis2 function of the vegan package (v.2.7-2) (42).

## RESULTS

Expression of *CREBRF*, as well as known adipose-specific genes (e.g. *ADIPOQ*), was observed in all samples. There was no difference in *CREBRF* expression among the three genotypes (p*_adj_*=0.89), but 22 significant (*p_adj_* < 0.1) DEGs were observed (**Table 2**). The distributions of gene expression within most of the DEGs were right-skewed, driven by 4-5 participants (different by DEG) with high expression (**Fig 3, Supp Fig 2,3**). Among the 22 DEGs, thirteen –*KRT1*, *SDC1*, *DSG1*, *A2ML1*, *AKAP6*, *IL1RL2*, *LYPD3*, *CAMSAP3*, *SCEL*, *S100A14*, *FLG*, *NMNAT2*, and *KLK1*—had a shared expression profile, with decreasing expression with each copy of the ‘A’ allele (**Fig 4**). Enriched gene ontology (GO) terms included: keratinocyte differentiation, epidermal cell differentiation, skin development, and epidermis development (**Fig 5, Supp Table 1**). In support of this finding, most DEGs were more highly expressed in skin compared to adipose in GTEx (**Fig 6**). Following deconvolution, all major adipose cell-types were detected in every sample (**Fig 7**). However, there were no differences in estimated cell-type proportions with the rs373863828 genotype (*p* > 0.05).

**Table 2:** Significance differentially expressed genes across rs373863828 genotypes.

| Symbol | Gene | Chromosome | ENSEMBL ID | BaseMean | p-value | Adjusted p-value* |
| --- | --- | --- | --- | --- | --- | --- |
| KRT1 | keratin 1 | 12 | ENSG00000167768 | 45.26 | 0.0000000 | 0.0002256 |
| KRT10 | keratin 10 | 17 | ENSG00000186395 | 1100.18 | 0.0000001 | 0.0008525 |
| SDC1 | syndecan 1 | 2 | ENSG00000115884 | 54.73 | 0.0000005 | 0.0038633 |
| DSG1 | desmoglein 1 | 18 | ENSG00000134760 | 17.49 | 0.0000010 | 0.0038633 |
| ATG9B | autophagy related 9B | 7 | ENSG00000181652 | 23.93 | 0.0000009 | 0.0038633 |
| SELENOM | selenoprotein M | 22 | ENSG00000198832 | 1346.97 | 0.0000011 | 0.0038633 |
| A2ML1 | alpha-2-macroglobulin like 1 | 12 | ENSG00000166535 | 8.54 | 0.0000014 | 0.0043863 |
| AKAP6 | A-kinase anchoring protein 6 | 14 | ENSG00000151320 | 37.06 | 0.0000018 | 0.0047770 |
| IGHV1-24 | immunoglobulin heavy variable 1-24 | 14 | ENSG00000211950 | 6.97 | 0.0000043 | 0.0102846 |
| CBX7 | chromobox 7 | 22 | ENSG00000100307 | 1096.97 | 0.0000156 | 0.0258814 |
| IL1RL2 | interleukin 1 receptor like 2 | 2 | ENSG00000115598 | 6.69 | 0.0000139 | 0.0258814 |
| LYPD3 | LY6/PLAUR domain containing 3 | 19 | ENSG00000124466 | 16.00 | 0.0000149 | 0.0258814 |
| KRT14 | keratin 14 | 17 | ENSG00000186847 | 210.58 | 0.0000143 | 0.0258814 |
| CAMSAP3 | calmodulin regulated spectrin associated protein family member 3 | 19 | ENSG00000076826 | 4.93 | 0.0000345 | 0.0532500 |
| SCEL | sciellin | 13 | ENSG00000136155 | 14.20 | 0.0000454 | 0.0652650 |
| S100A14 | S100 calcium binding protein A14 | 1 | ENSG00000189334 | 8.81 | 0.0000509 | 0.0687153 |
| FLG | filaggrin | 1 | ENSG00000143631 | 16.93 | 0.0000607 | 0.0770471 |
| TMEM109 | transmembrane protein 109 | 11 | ENSG00000110108 | 2616.90 | 0.0000925 | 0.0951046 |
| NMNAT2 | nicotinamide nucleotide adenylyltransferase 2 | 1 | ENSG00000157064 | 279.92 | 0.0000912 | 0.0951046 |
| KLK1 | kallikrein 1 | 19 | ENSG00000167748 | 3.87 | 0.0000885 | 0.0951046 |
| LINC00311 | long intergenic non-protein coding RNA 311 | 16 | ENSG00000179219 | 5.92 | 0.0000813 | 0.0951046 |
| STRA6 | signaling receptor and transporter of retinol STRA6 | 15 | ENSG00000137868 | 6.08 | 0.0001019 | 0.0999472 |

\* Benjamini-Hochberg corrected

**Figure 3.**
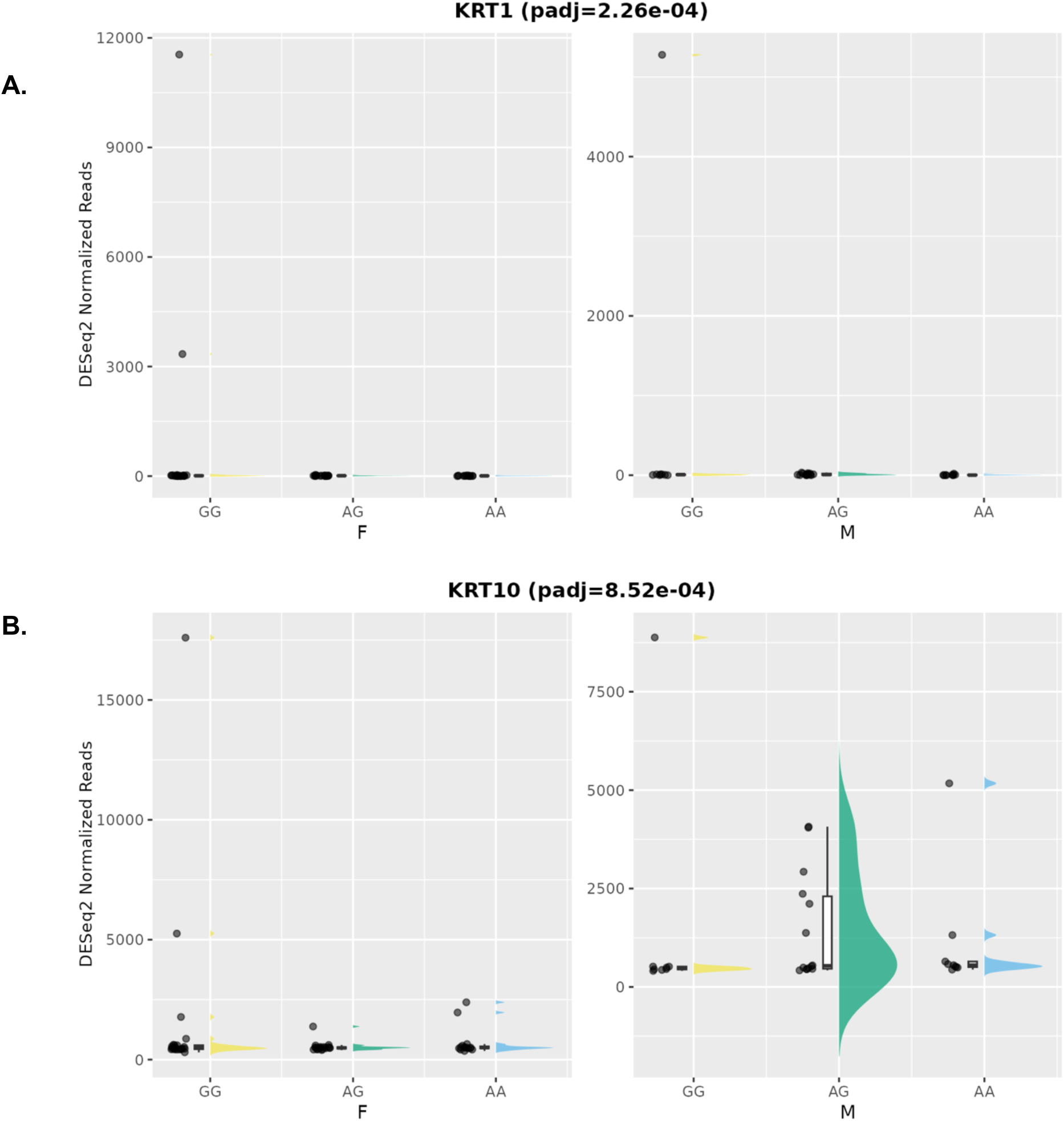
Rain cloud plots of expression of the top two differentially expressed genes, (A) *KRT1* and (B) *KRT10,* stratified by sex and rs373863828 genotype. Sex is indicated as F for female and M for male along the x-axis. Expression as measured by DESeq2 normalized reads. Quartiles and median of DESeq2 normalized reads for respective combination of rs373863828 genotype and sex are represented by the box plots, with individual samples represented by jittered points. Half-violin plots illustrate the shape of the distribution. Rain cloud plots for the remaining 20 DEGs available in **Supplemental** Figure 3.

**Figure 4.**
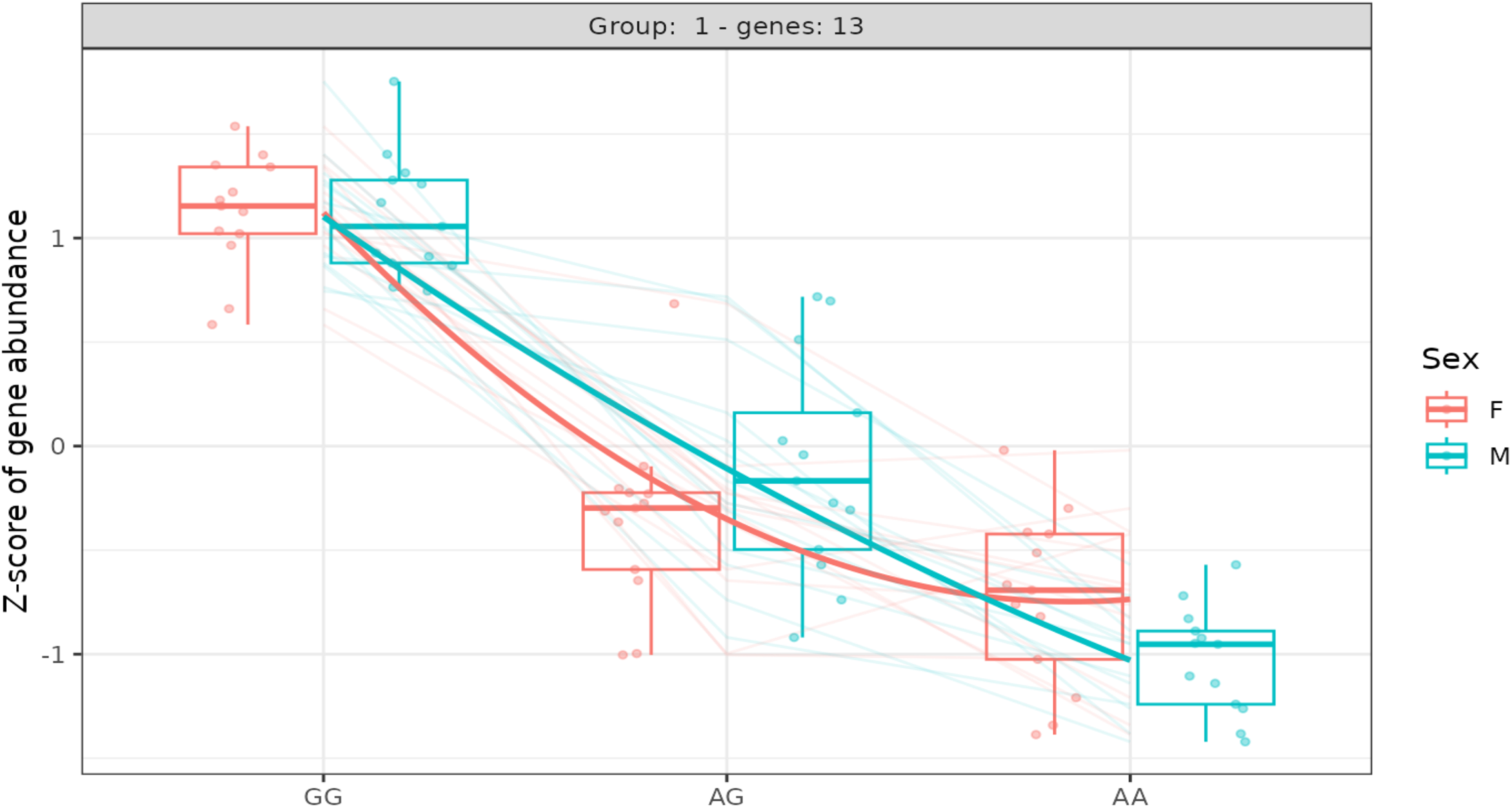
Shared profile of 13 differentially expressed genes across the three rs373863828 genotypes. Expression of *KRT1, SDC1, DSG1, A2ML1, AKAP6, IL1RL2, LYPD3, CAMSAP3, SCEL, S100A14, FLG, NMNAT2,* and *KLK1* decreases with each copy of the ‘A’ allele.

**Figure 5.**
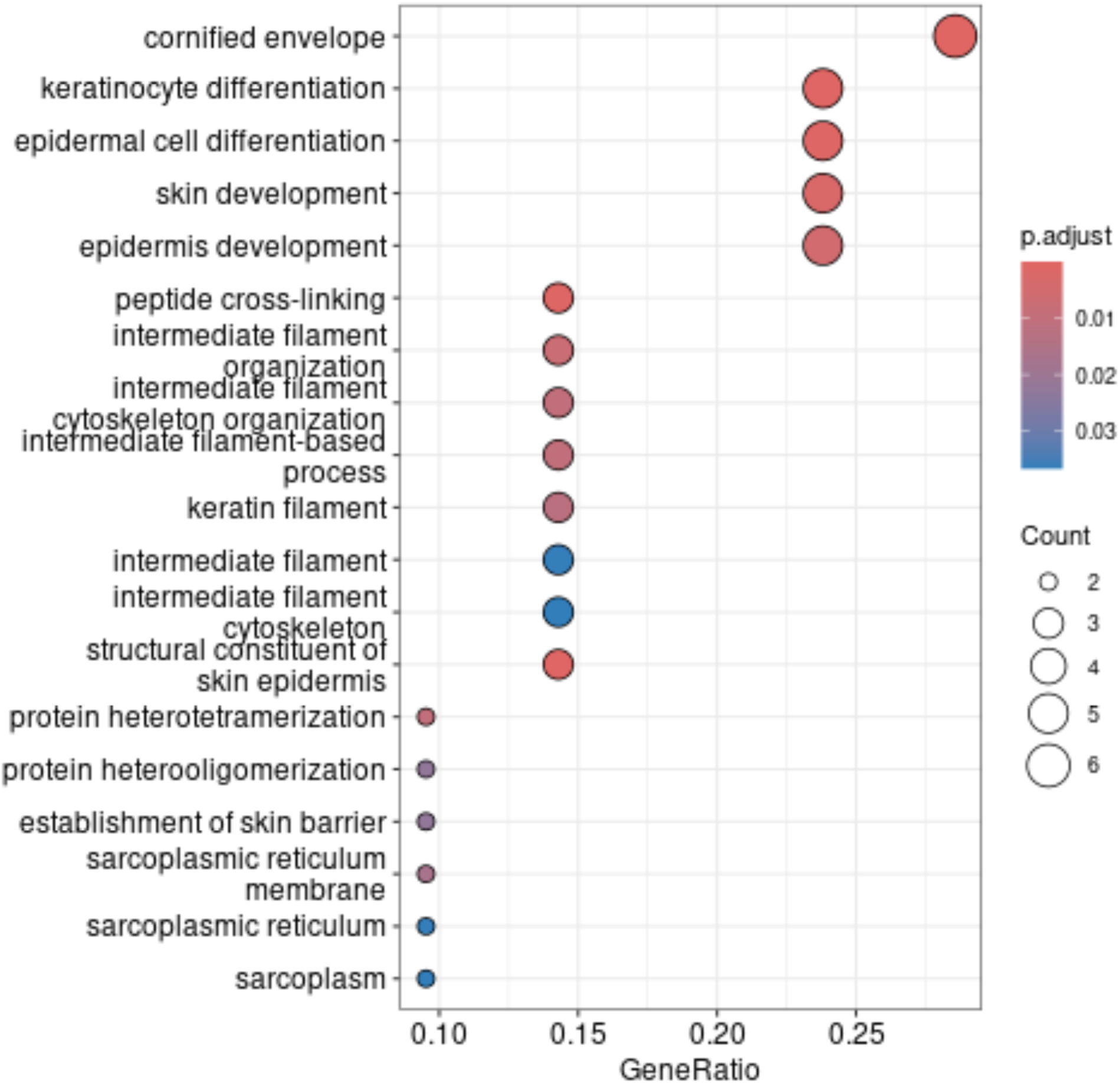
Enrichment analysis. indicates that significantly differentially expressed genes are enriched for skin-related gene ontology (GO) terms. clusterProfiler was used to identify enriched GO terms for molecular function, biological process, and cellular component.

**Figure 6.**
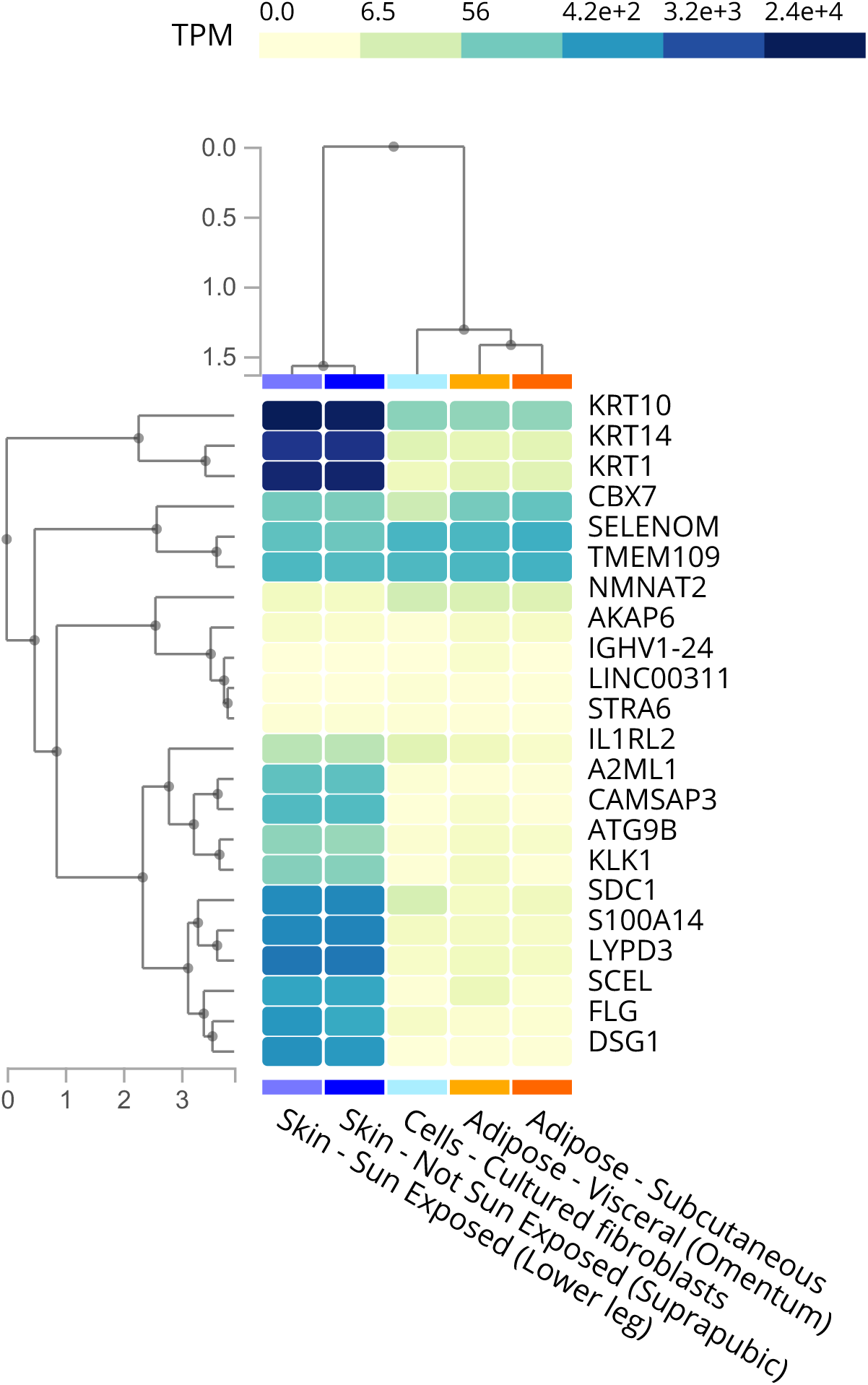
Expression of significant differentially expressed genes (*p_adj_* < 0.10) in adipose and skin per the Adult Genotype Tissue Expression (GTEx) portal [accessed 13 August 2024].

**Figure 7.**
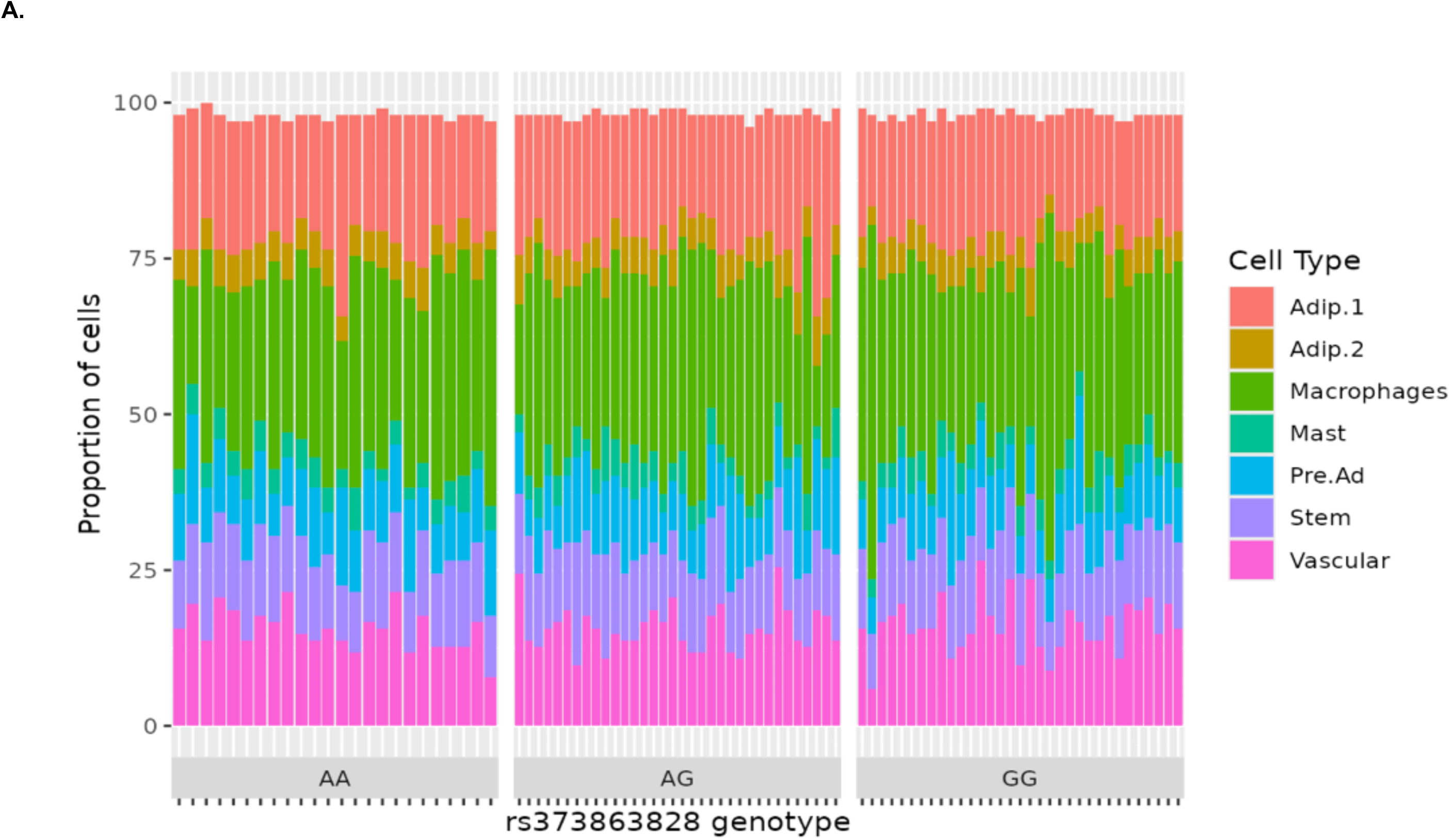
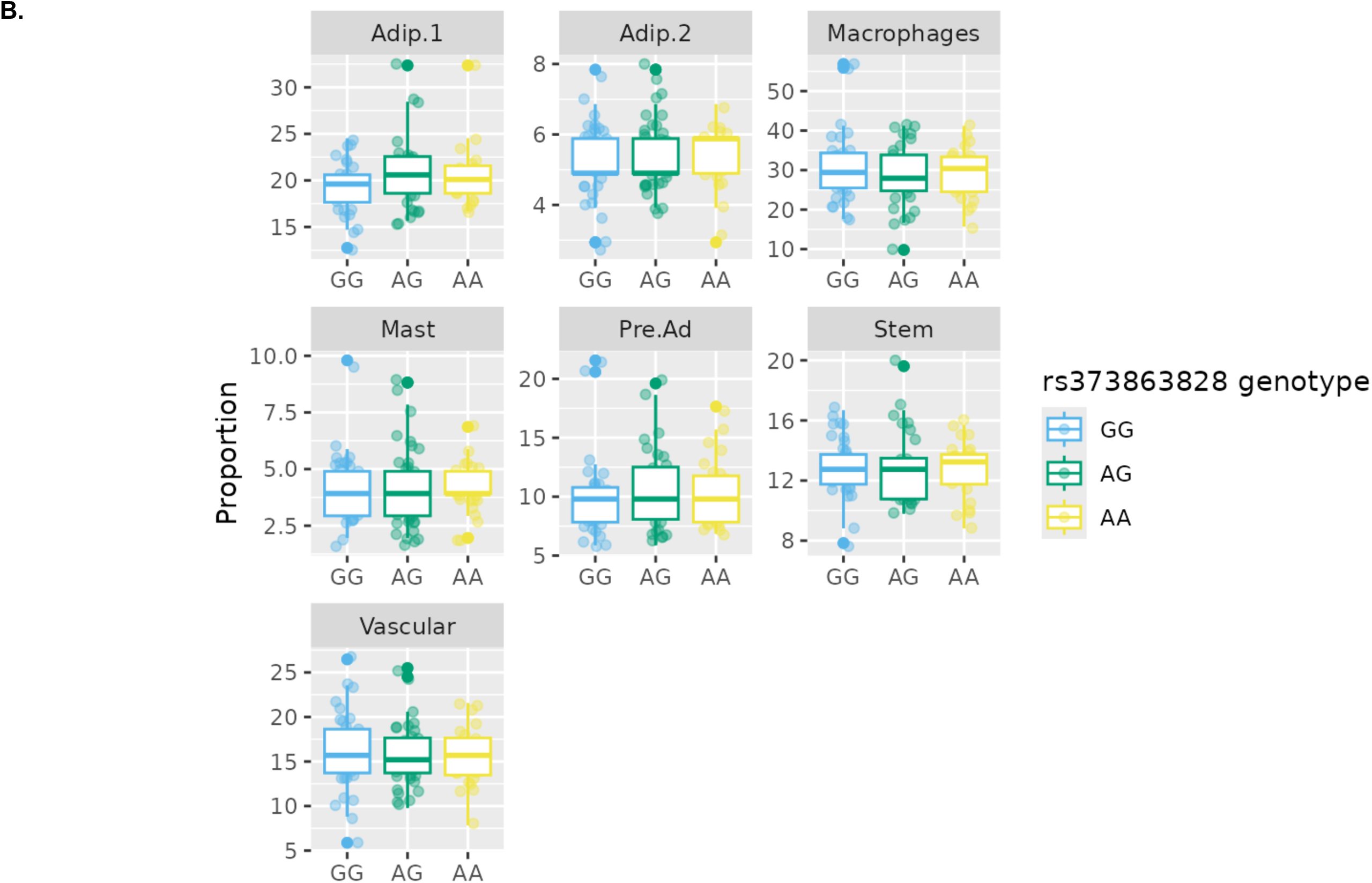
Deconvolution-derived cell type proportions are similar across rs373863828 genotypes. PERMANOVA shows no significant differences (*p* > 0.05) among cell types by rs373863828 genotype. **(A)** Stacked bar chart of cell type proportions grouped by genotype. **(B)** Boxplots of each cell type by genotype.

## DISCUSSION

Here we describe the first study, to our knowledge, of the effect of a population-specific variant, *CREBRF* rs373863828 (p.Arg457Gln), on gene expression in human adipose tissue. Our results indicate that, despite expectations that differences may be observed in this metabolically active tissue, few genes are differentially expressed in adipose tissue due to rs373863828 genotype. As expected, some of the DEGs, particularly those that are more highly expressed in adipose tissue, have described roles in adipose and metabolism. For example, polycomb protein Chromobox 7 (*CBX7, p_adj_*=0.026) is a negative regulator of adipogenesis, diminishing differentiation and lipid accumulation when overexpressed in 3T3-L1 cells or adipose-derived stem cells from *Cbx-7* overexpressing mice (43). Selenoprotein M (*SELENOM*; *p_adj_*= 0.004) promotes adipogenesis in 3T3-L1 cells, and *SelenoM* knock-out mice fed high-fat diet have poorer metabolic profiles compared to their control littermates (44).

Some of the DEGs that were identified are enriched for several skin-related GO terms and are highly expressed in skin (**Figs. 5,6**). For example, multiple keratins, including *KRT1* (*p_adj_*= 0.0002) and *KRT10* (*p_adj_*= 0.0009), were among the top DEGs (**Table 2**). *KRT1* and *KRT10* are highly expressed in differentiating keratinocytes and hair follicles (45). However, adipose and skin do have an established relationship, and a distinct layer of dermal-associated adipocytes (dermal white adipose tissue, dWAT) exists between the reticular dermis and sWAT (46,47). Interestingly, *Sdc-1* deficient mice have significantly decreased dWAT and are smaller compared to wild-type mice (48). Furthermore, obesity and elevated levels of sWAT are risk factors for a variety of dermal conditions via delayed wound healing, altered skin barrier, and decreased dermal elasticity, conditions also observed in T2D (49–51).

The small number of DEGs identified in this study may be expected. The variant is a single base substitution located in an exon (p.R457Q). Most regulatory variants are located within non-coding regions of the genome, and while coding variants have been reported to alter gene expression, they primarily do so by introducing an alternative splice site or premature stop codon, neither of which are consequences of rs373863828 (52–54). However, other explanations may exist.

Adipose tissue is heterogeneous, comprised of mature and immature adipocytes, progenitor cells, endothelial cells, and macrophages (55–57). If gene expression is altered in only a subset of these cell populations, bulk sequencing is unlikely to detect cell-type specific changes (55,58). Similarly, *CREBRF* may be exerting its effects through other adipose depots relevant for insulin resistance.

A limitation of this study is that participants self-reported fasting prior to the biopsy, so the length of and adherence to fasting requirements could not be confirmed. This is particularly relevant given that *CREBRF* expression is upregulated during nutrient starvation - in mice, 3T3-L1 cells, HeLa cells, and SAOS-2 cells - and quickly rebounds following the reintroduction of nutrients (1,59); this response is more pronounced in cells containing the variant or overexpressing *CREBRF*. The length of fasting for study participants may have been insufficient to recapitulate the starvation conditions driving the effects of rs373863828 genotype and increased *CREBRF* expression seen in mouse and cellular models, especially for individuals with high BMI who likely have large energy stores. Further, rs373863828 is associated with a decreased odds of T2D despite its association with higher mean BMI. We recruited healthy individuals and excluded those with known major cardiometabolic disease to determine baseline effects of the variant unrelated to any pathophysiology, and therefore are unable to detect any disease-specific DGE that may exist (58).

Mice overexpressing *Crebrf* are overweight compared to wild-type mice (unpublished), and 3T3-L1 cells overexpressing human *CREBRF* accumulate more lipids and undergo adipogenesis faster than wild-type cells (1). However, Kanshana and Metcalfe report that variant knock-in mouse models fail to recapitulate the human phenotype, despite high conservation of the gene between the two species (60,61). It is therefore possible that the effects of rs373863828 genotype on *CREBRF* activity and associated phenotypes are modulated by organism-specific regulation and/or epistatic interactions.

Despite the minimal impact of this variant on gene expression in this study, the substantial impact of *CREBRF* on cardiometabolic phenotypes has been replicated in multiple cohorts and requires additional investigation to determine how it so strongly influences these traits (1–8,13) and how those mechanisms of action might be leveraged for behavioral or pharmaceutical prevention or treatment. Future work should assess the impact of rs373863828 genotype in other tissue types, while single-cell approaches may provide a more refined picture of this variant’s impact on gene expression on specific cell populations (62).

## ACKNOWLEDGEMENTS AND FUNDING

We would like to acknowledge and express our sincere gratitude to the research participants. We also thank the Samoan government, especially the Ministry of Health; the Ministry of Women, Community, and Social Development; the Office of the Prime Minister; and the Samoa Bureau of Statistics for their support of this work. We thank Dr. Alec Ekeroma and Alec’s Home Away (Apia, Samoa) for providing clinical space to perform the adipose biopsies.

This work was supported by the US National Institutes of Health (NIH) NHLBI R01HL093093 (Principal Investigator [PI]: Stephen T. McGarvey, Brown University), NCI T32CA175294 (PI: John M. Kirkwood, University of Pittsburgh), NIDDK T32DK007052 (PI: Robert M. O’Doherty, University of Pittsburgh), and NCATS UL1TR001857 (PI: Steven E Reis, University of Pittsburgh). ACR was supported by the US NIH and Fogarty International Center (Global Health Equity Scholars program D43TW010540). This project used the University of Pittsburgh HSCRF Genomics Research Core, RRID: SCR_018301 for Agilent TapeStation System.

## Data availability statement

Data for this study are unavailable for public use as a criterion of the participant consent. Please contact the corresponding author for data access requests. Code available upon request.

