## Supplemental Figures for "*CREBRF* rs373863828 minimally regulates gene expression in subcutaneous adipose tissue of Samoan adults"

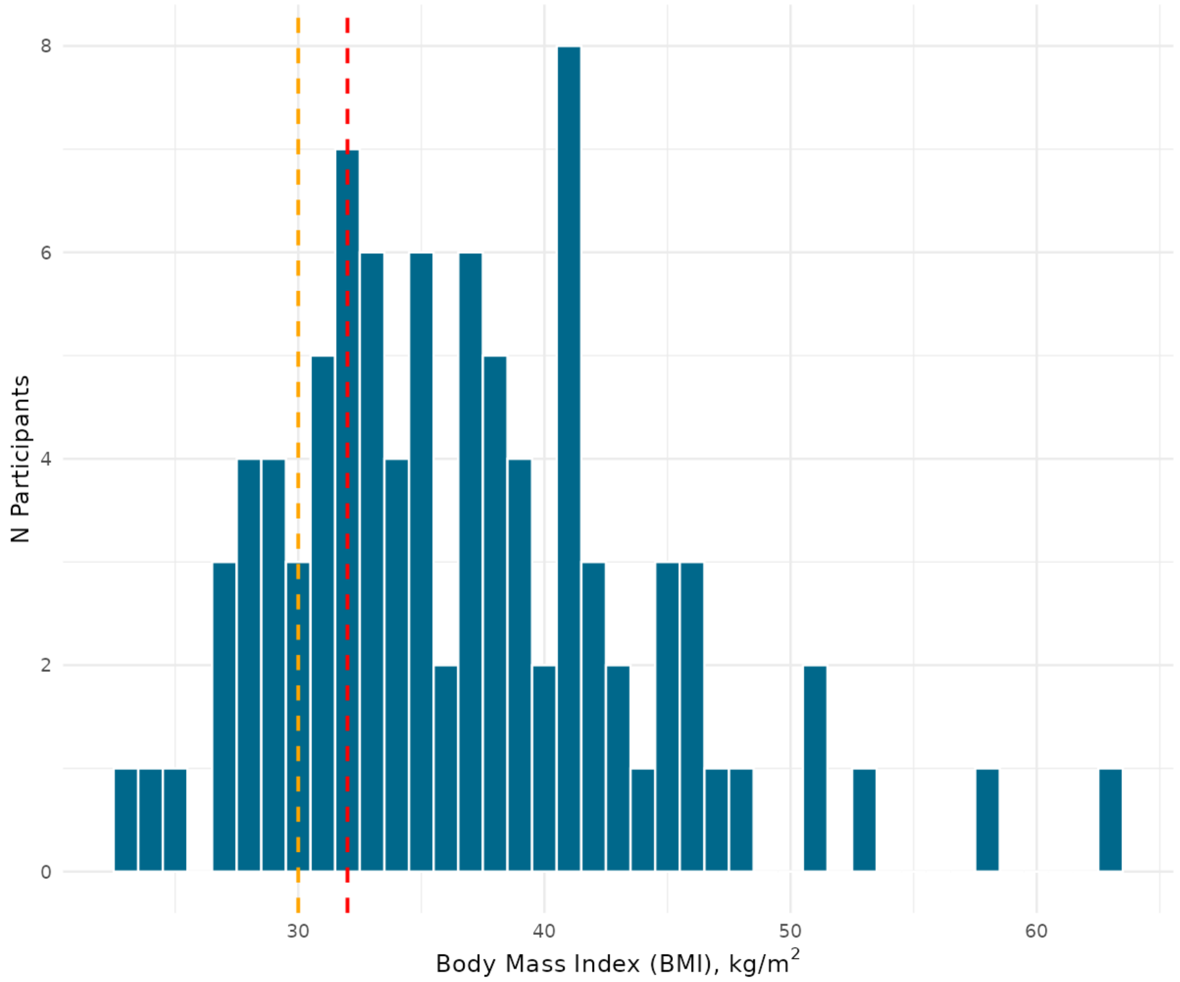


**Supplemental Figure 1. Body mass distribution of participants (n=91).** Reference lines indicate Polynesian cutoffs for overweight (BMI > 30, orange) and obese (BMI >32, red) weight classes.

A.

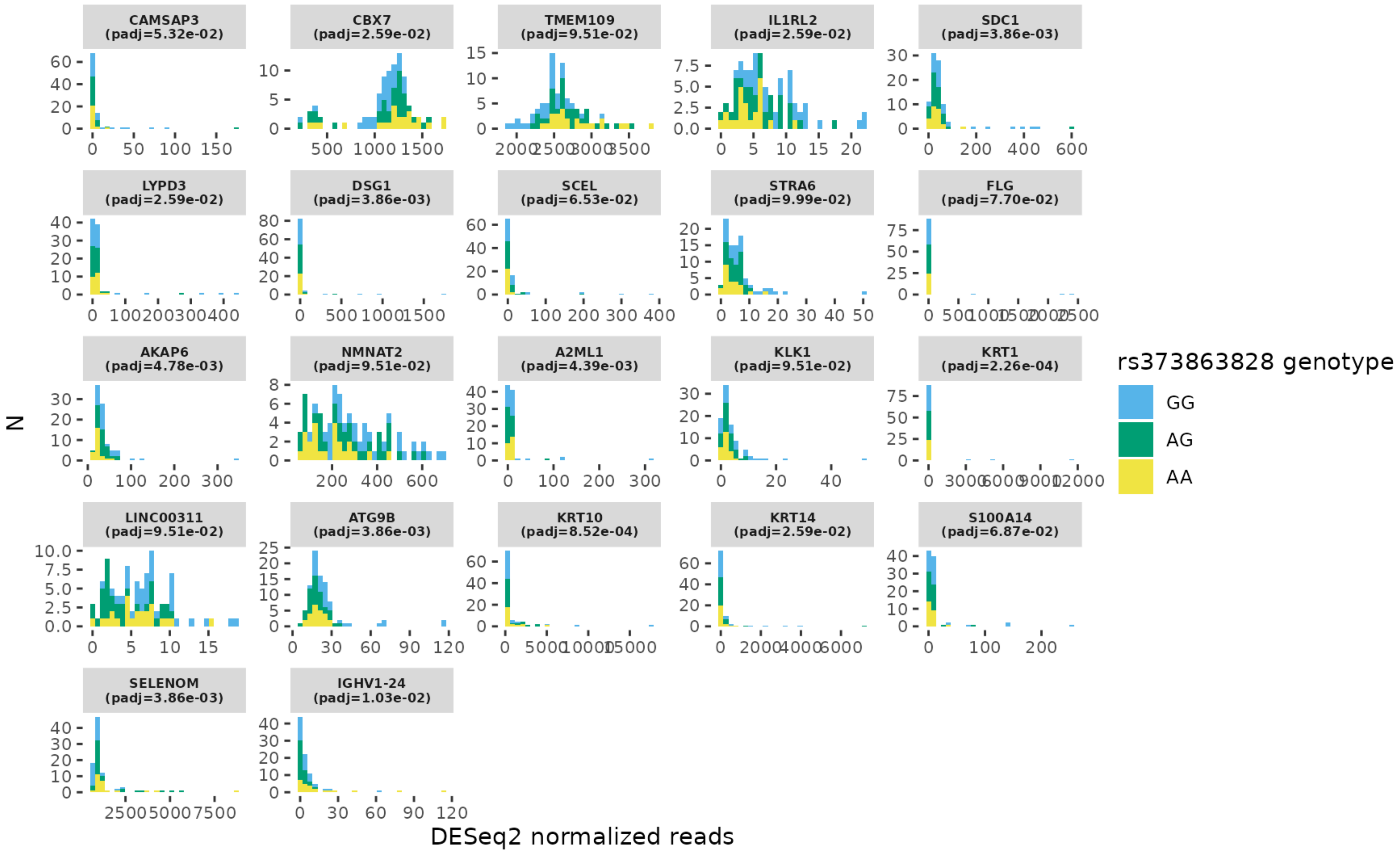

CAMSAP3 (padj=5.32e-02)
CBX7 (padj=2.59e-02)
TMEM109 (padj=9.51e-02)
IL1RL2 (padj=2.59e-02)
SDC1 (padj=3.86e-03)
LYPD3 (padj=2.59e-02)
DSG1 (padj=3.86e-03)
SCEL (padj=6.53e-02)
STRA6 (padj=9.99e-02)
FLG (padj=7.70e-02)
AKAP6 (padj=4.78e-03)
NMNAT2 (padj=9.51e-02)
A2ML1 (padj=4.39e-03)
KLK1 (padj=9.51e-02)
KRT1 (padj=2.26e-04)
LINC00311 (padj=9.51e-02)
ATG9B (padj=3.86e-03)
KRT10 (padj=8.52e-04)
KRT14 (padj=2.59e-02)
S100A14 (padj=6.87e-02)
SELENOM (padj=3.86e-03)
IGHV1-24 (padj=1.03e-02)
N
DESeq2 normalized reads
rs373863828 genotype
GG
AG
AA

**B.**

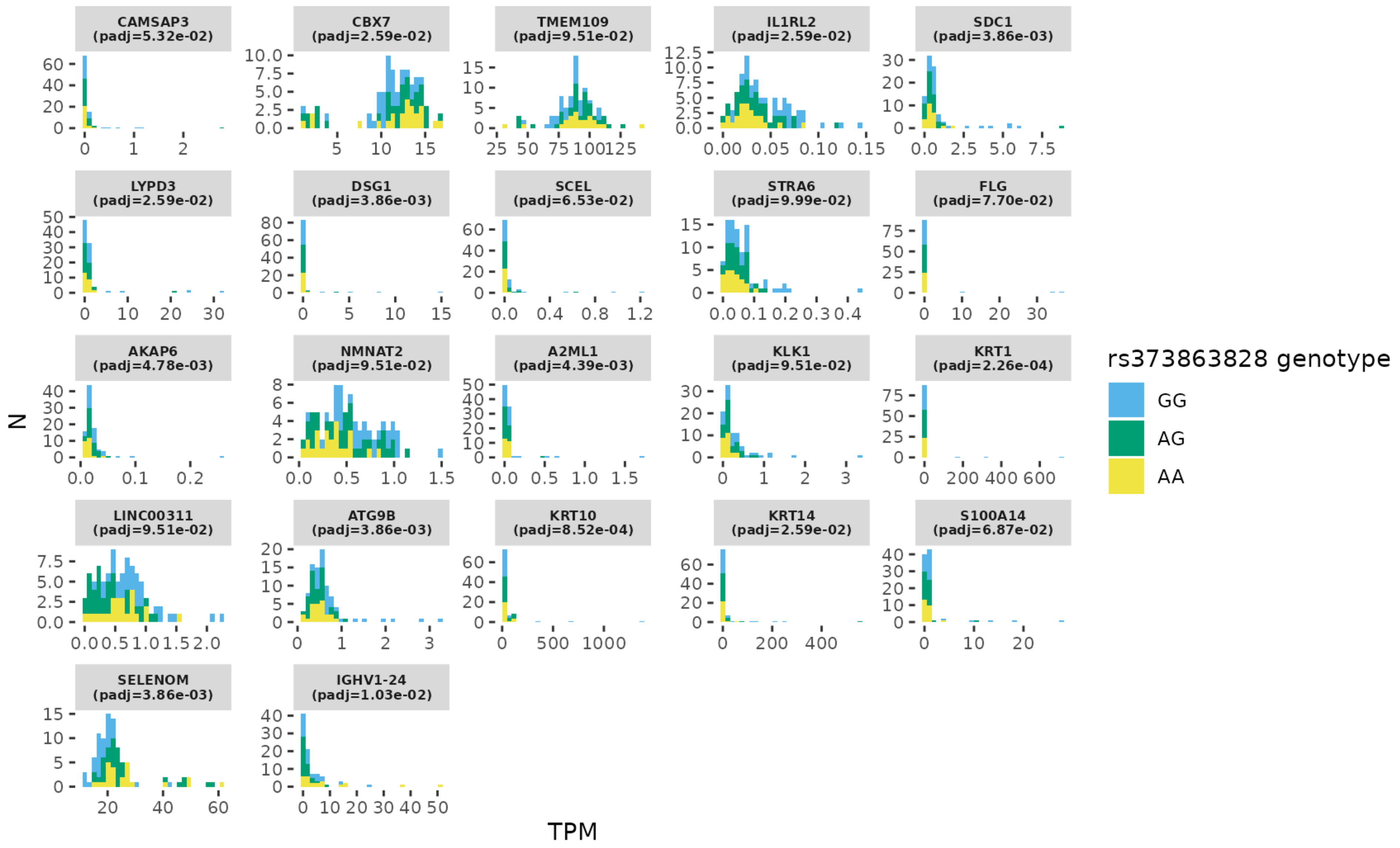


**Supplemental Figure 2. Distribution of (A) DESeq2 normalized reads and (B) transcripts per million (TPM) for significant differentially expressed genes (DEGs).** For **(B)**, gene lengths were obtained using the AnnotationHub package (v.3.14.0; hub: AH73881), and TPM was calculated using the convertCounts function of the DGEobj.utils package (v.1.0.6).

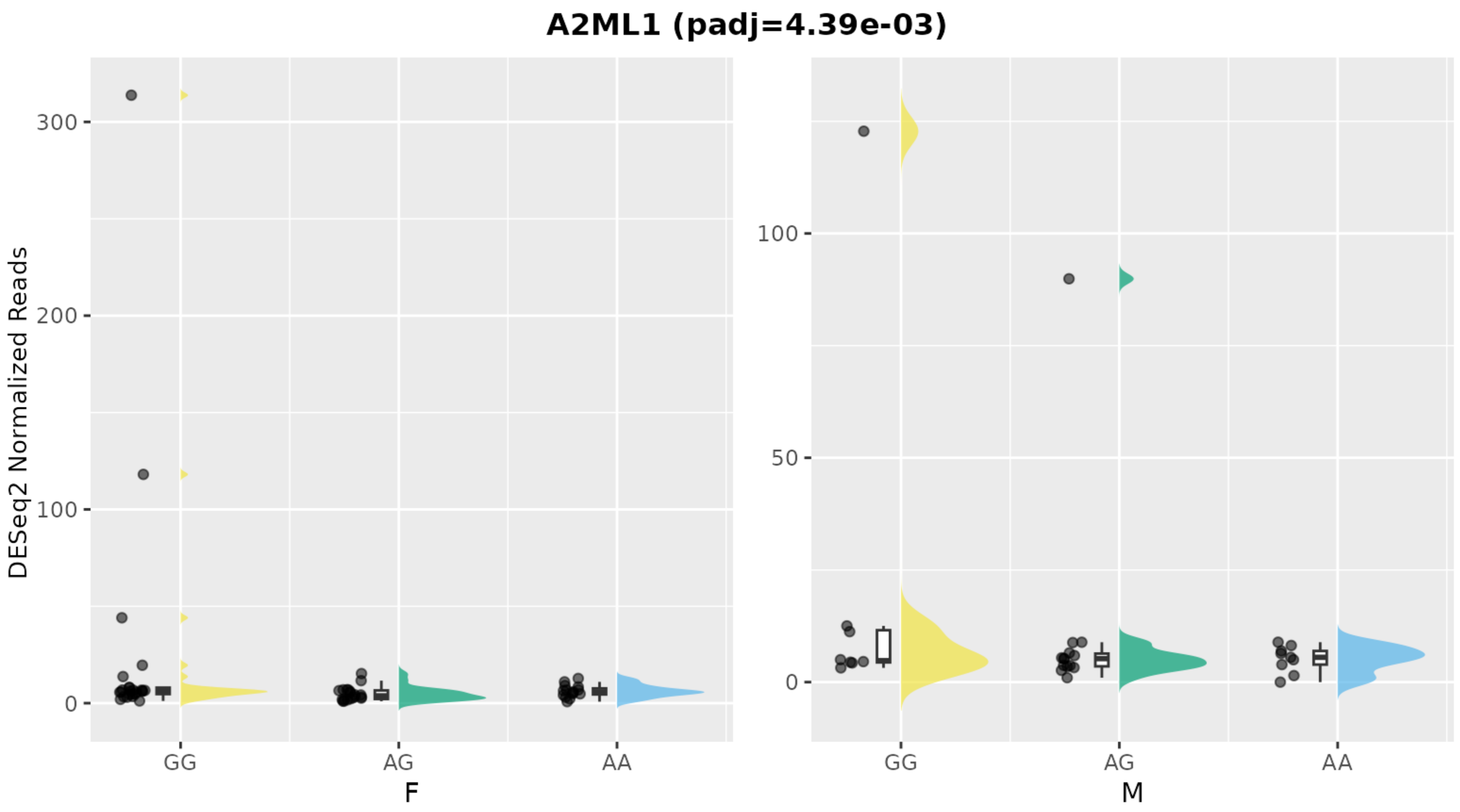

A2ML1 (padj=4.39e-03)
DESeq2 Normalized Reads
300
200
100
0
GG
AG
AA
F
100
50
0
GG
AG
AA
M

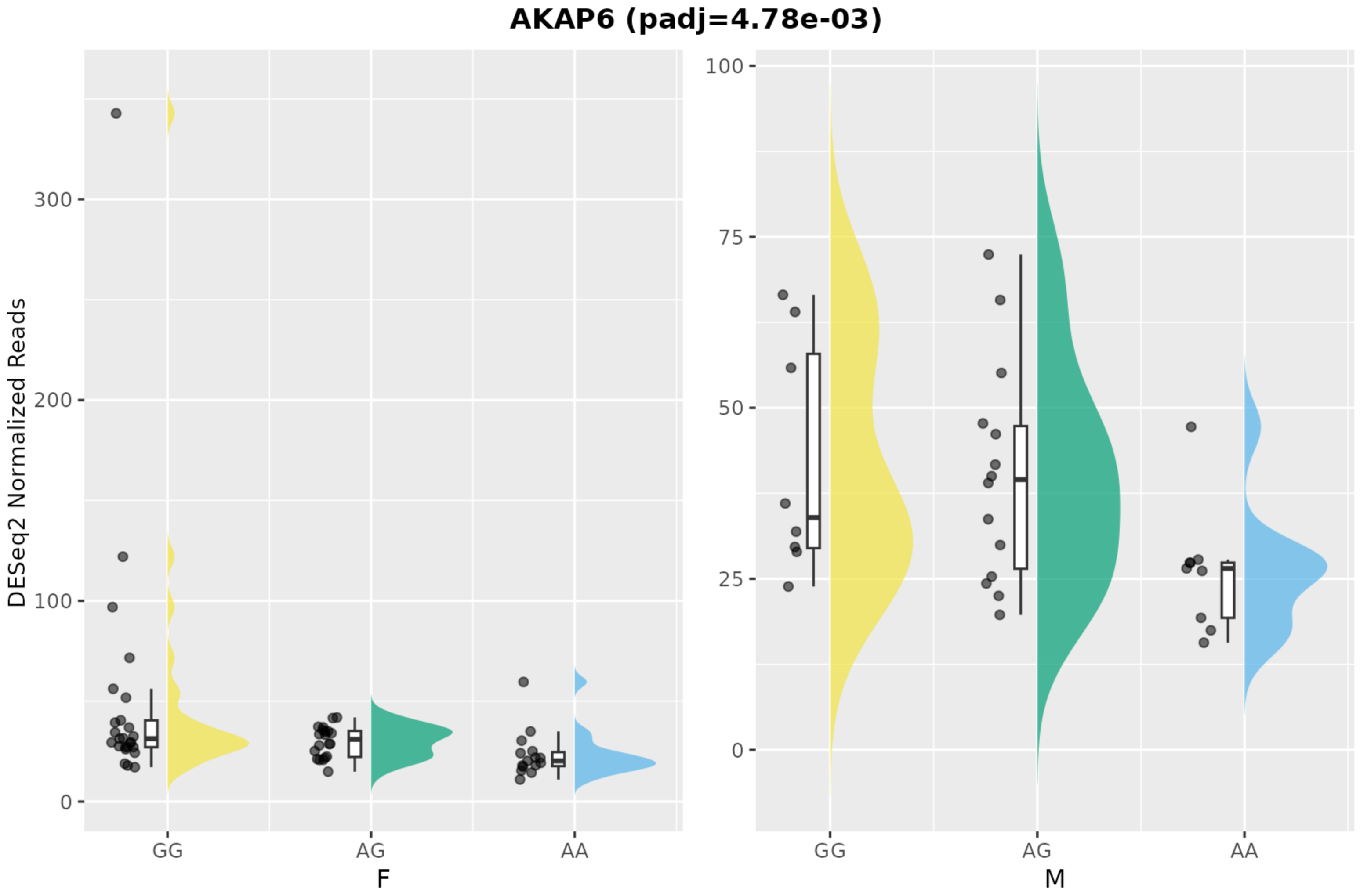

AKAP6 (padj=4.78e-03)
DESeq2 Normalized Reads
300
200
100
0
GG
AG
AA
F
100
75
50
25
0
GG
AG
AA
M

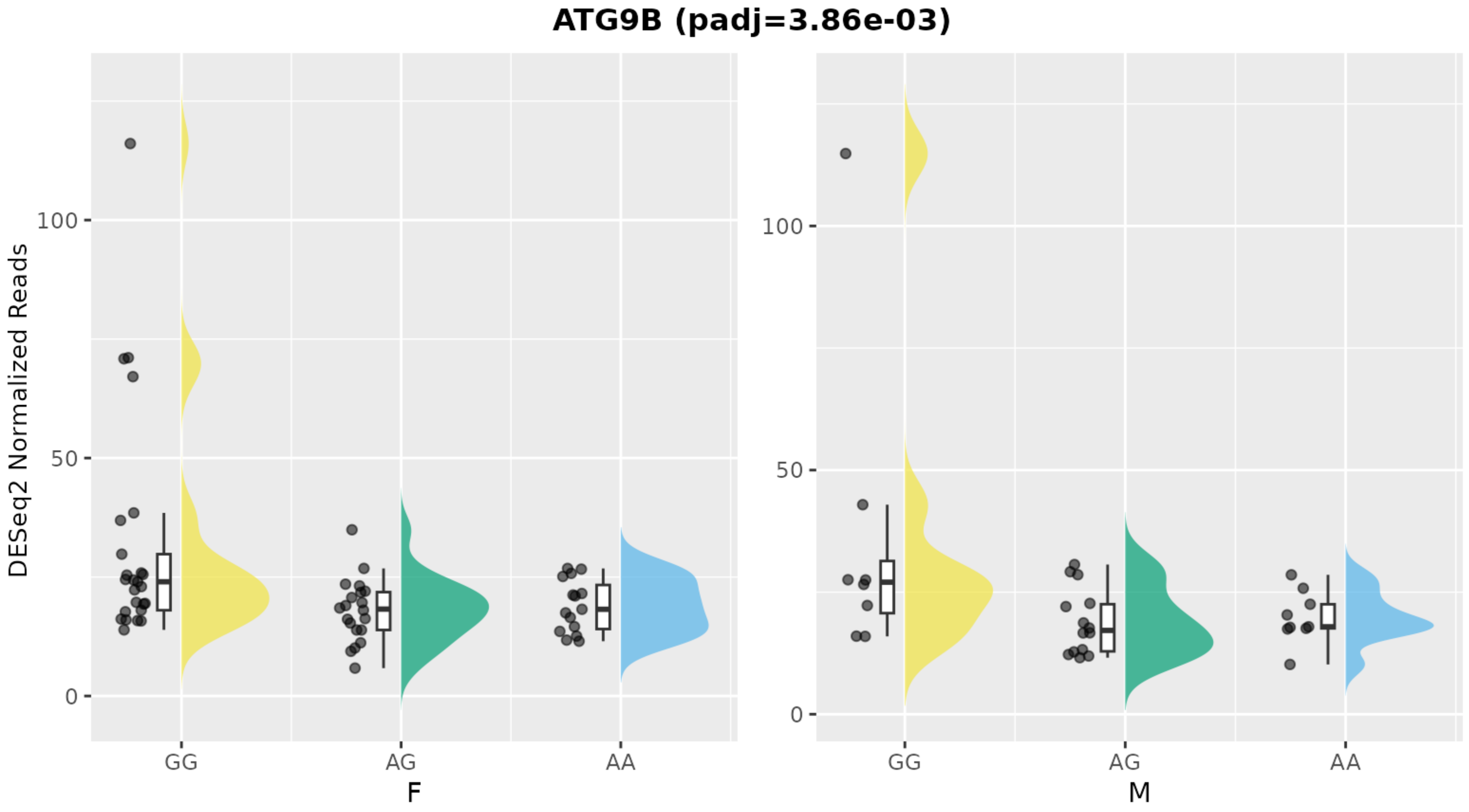
ATG9B (padj=3.86e-03)
DESeq2 Normalized Reads
100
50
0
GG
AG
AA
F
M

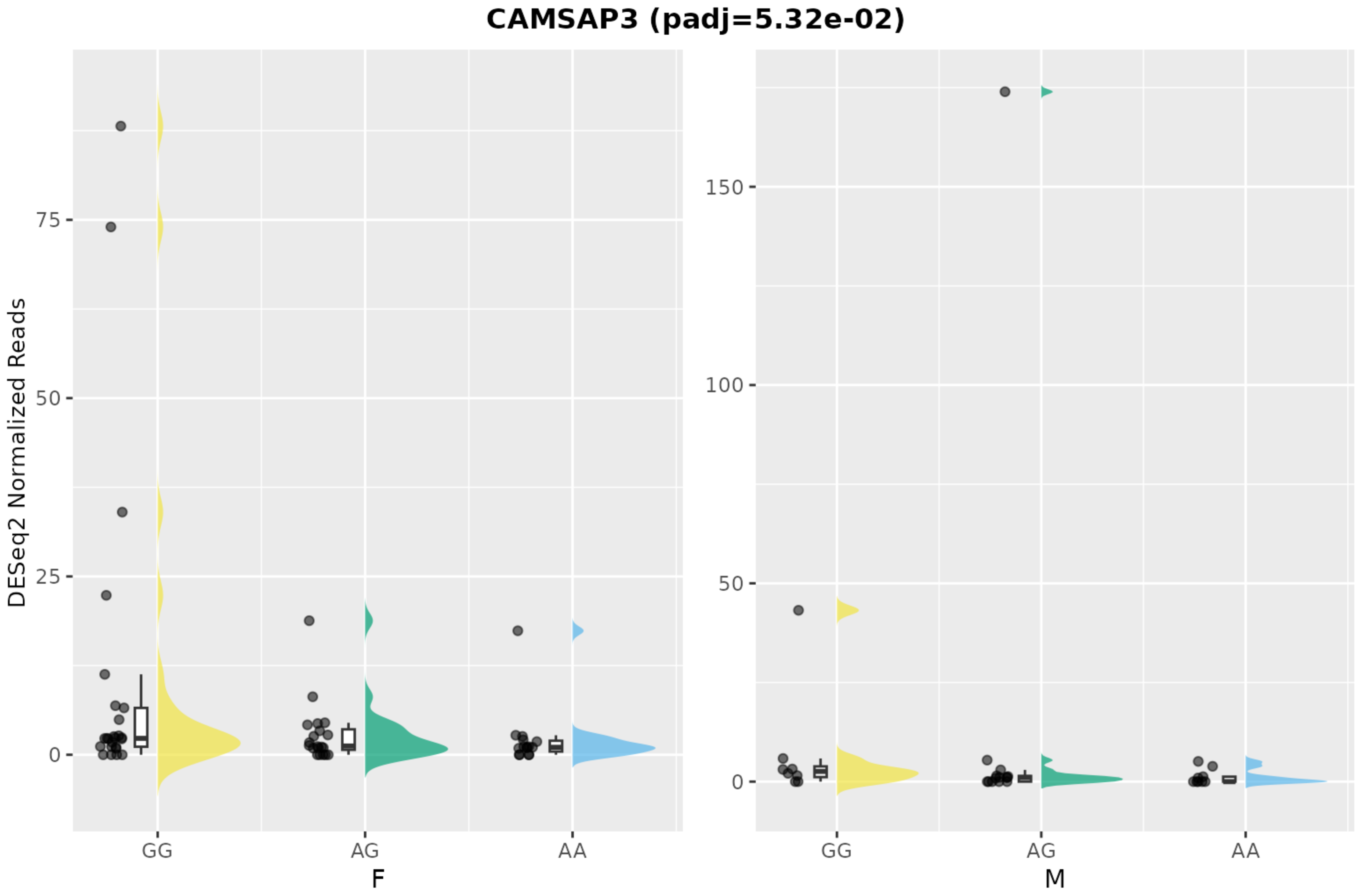
CAMSAP3 (padj=5.32e-02)
DESeq2 Normalized Reads
75
50
25
0
150
100
50
0
GG
AG
AA
F
GG
AG
AA
M

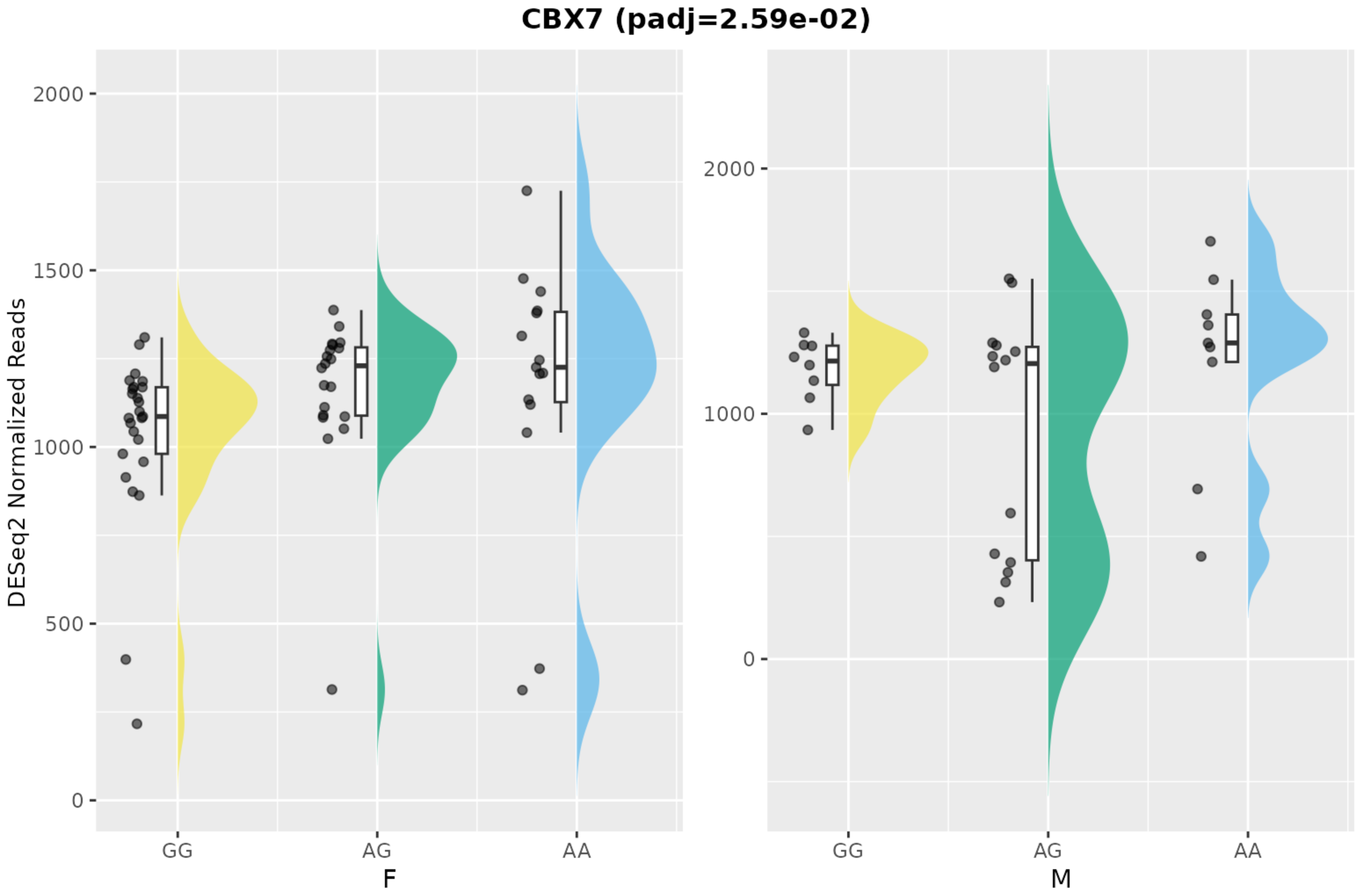

CBX7 (padj=2.59e-02)
DESeq2 Normalized Reads
2000
1500
1000
500
0
GG
AG
AA
F
M

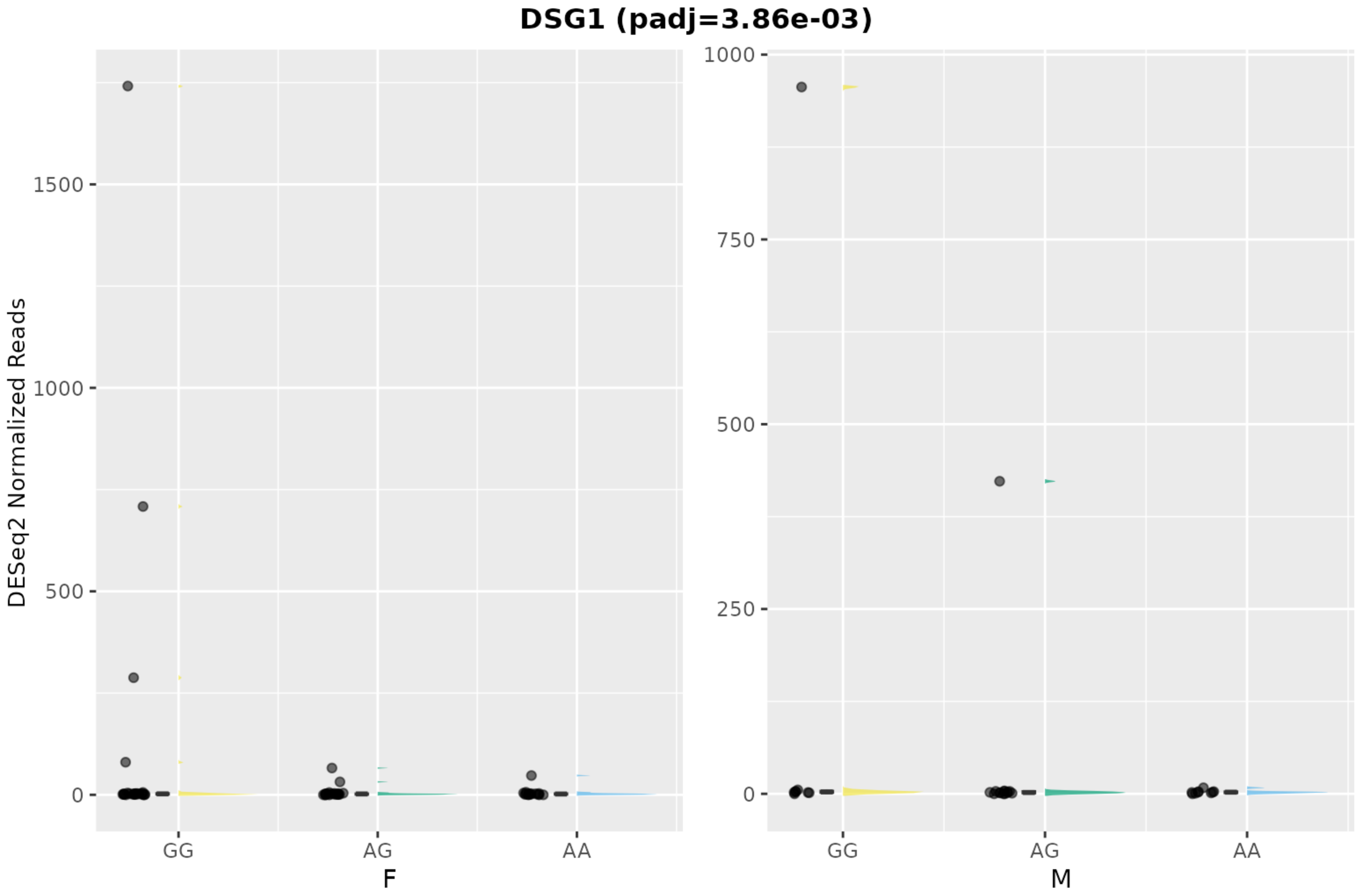
DSG1 (padj=3.86e-03)
DESeq2 Normalized Reads
1500
1000
500
0
GG
AG
AA
F
1000
750
500
250
0
GG
AG
AA
M

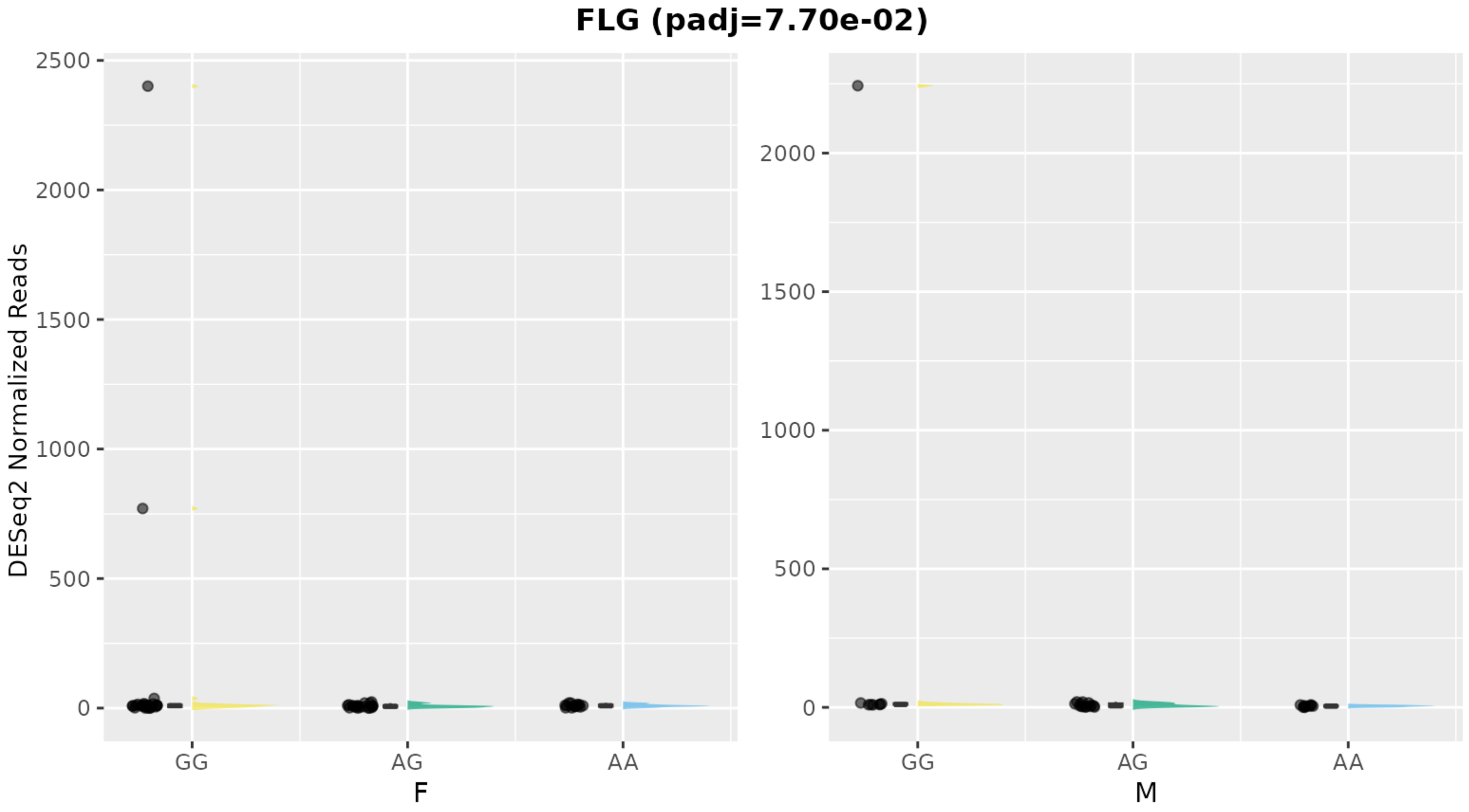

FLG (padj=7.70e-02)
DESeq2 Normalized Reads
2500
2000
1500
1000
500
0
GG
AG
AA
F
M

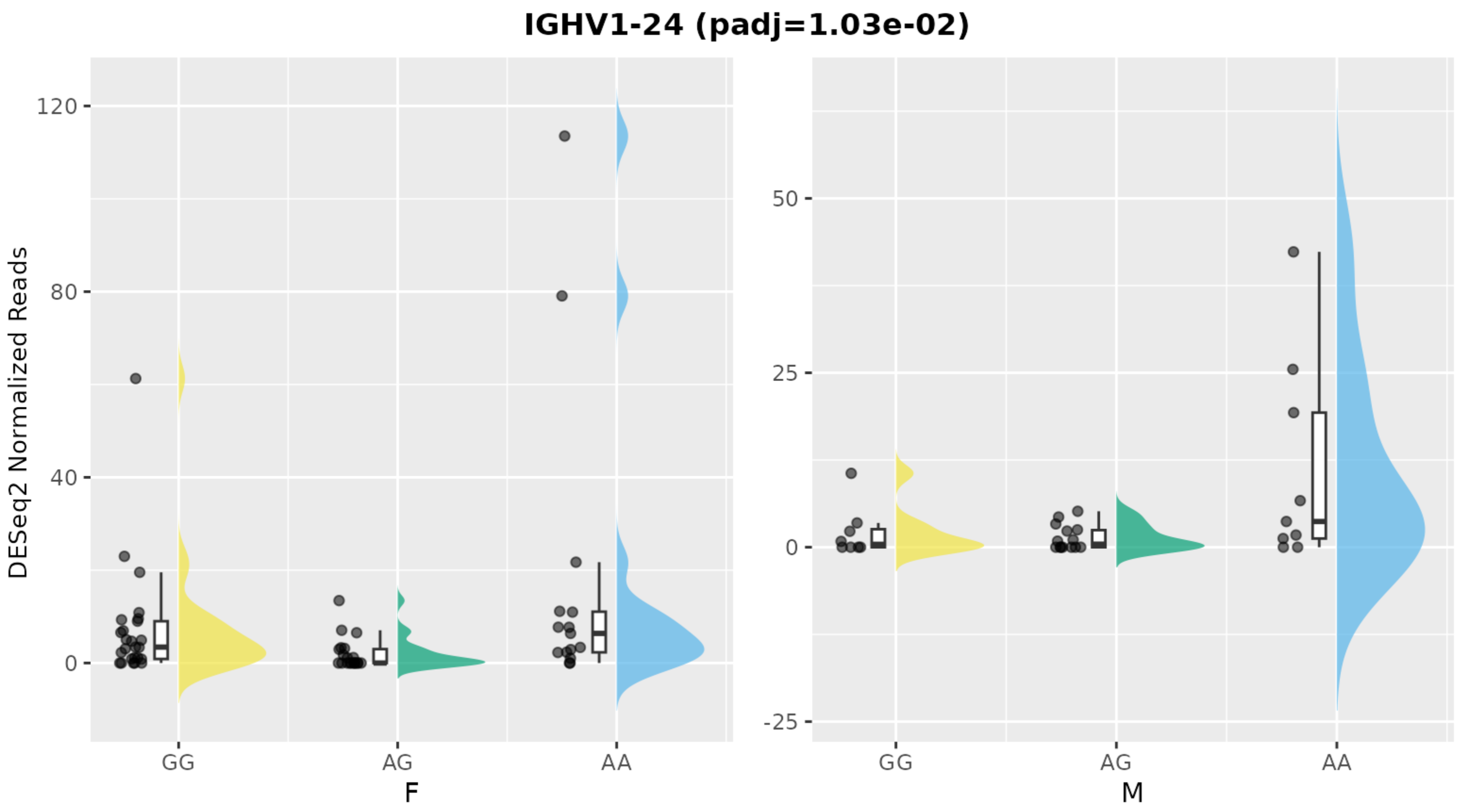
IGHV1-24 (padj=1.03e-02)
DESeq2 Normalized Reads
120
80
40
0
GG
AG
AA
F
50
25
0
-25
GG
AG
AA
M

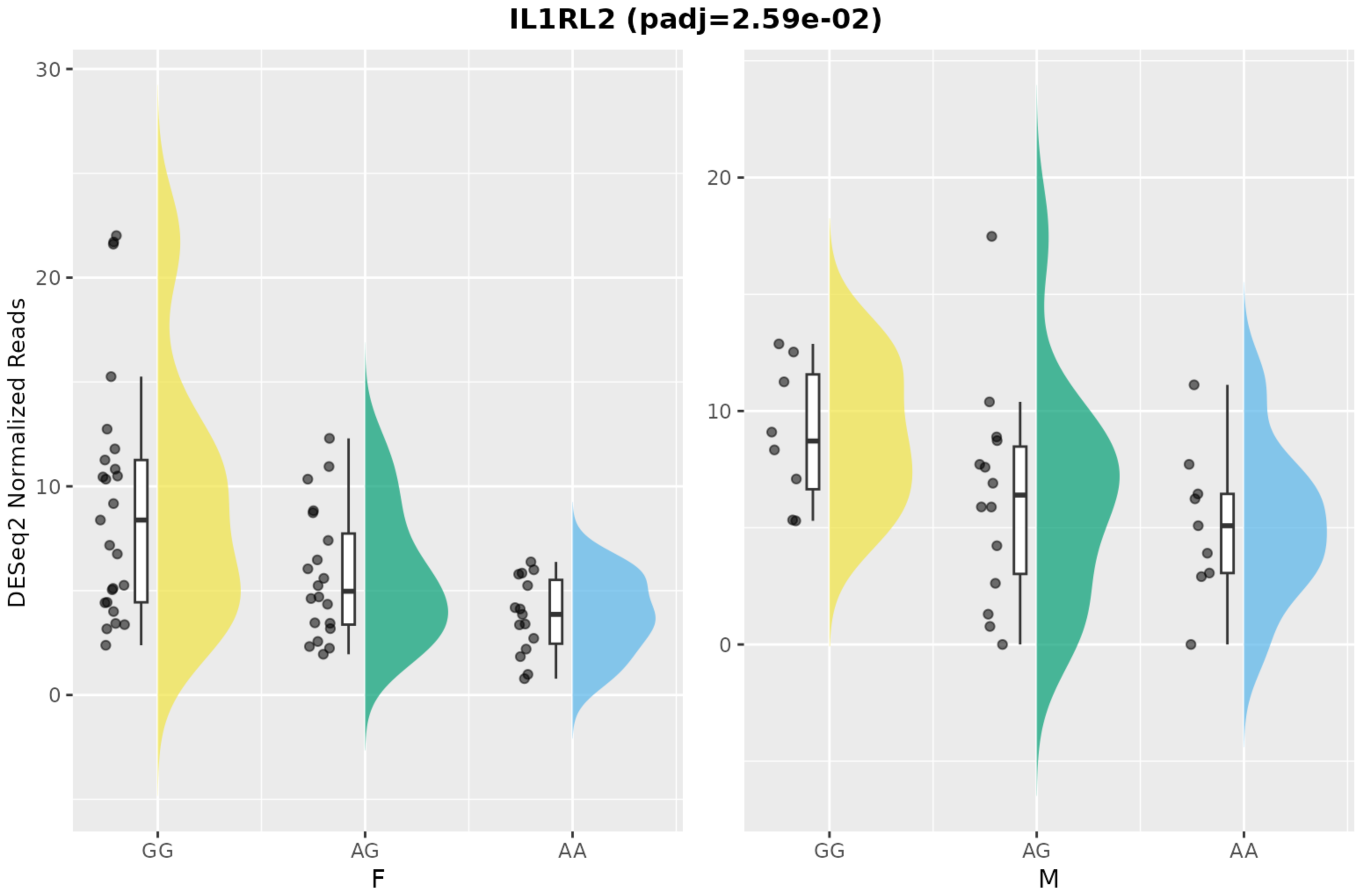

IL1RL2 (padj=2.59e-02)
DESeq2 Normalized Reads
30
20
10
0
GG
AG
AA
F
20
10
0
GG
AG
AA
M

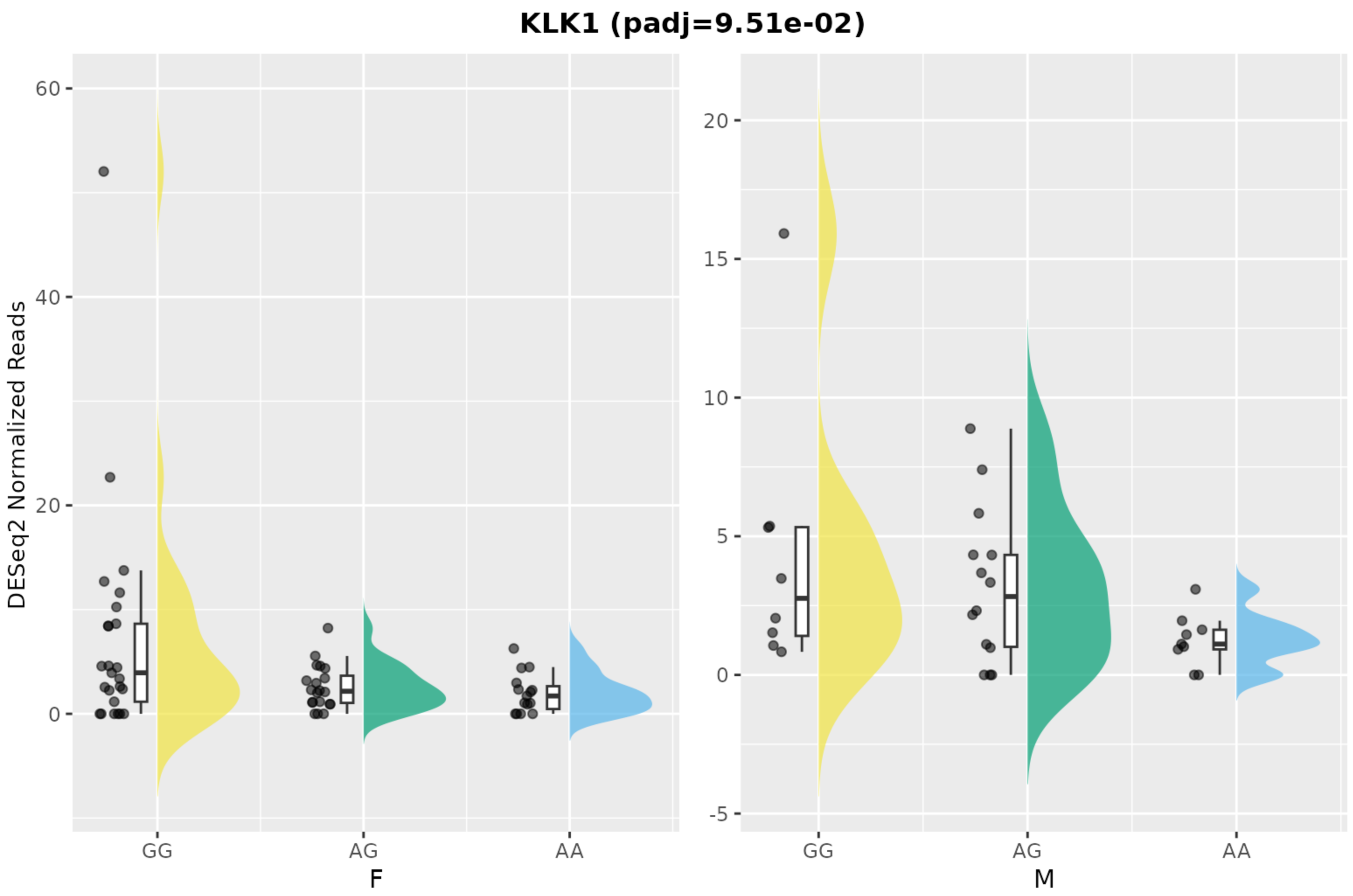
KLK1 (padj=9.51e-02)
DESeq2 Normalized Reads
60
40
20
0
GG
AG
AA
F
20
15
10
5
0
-5
GG
AG
AA
M

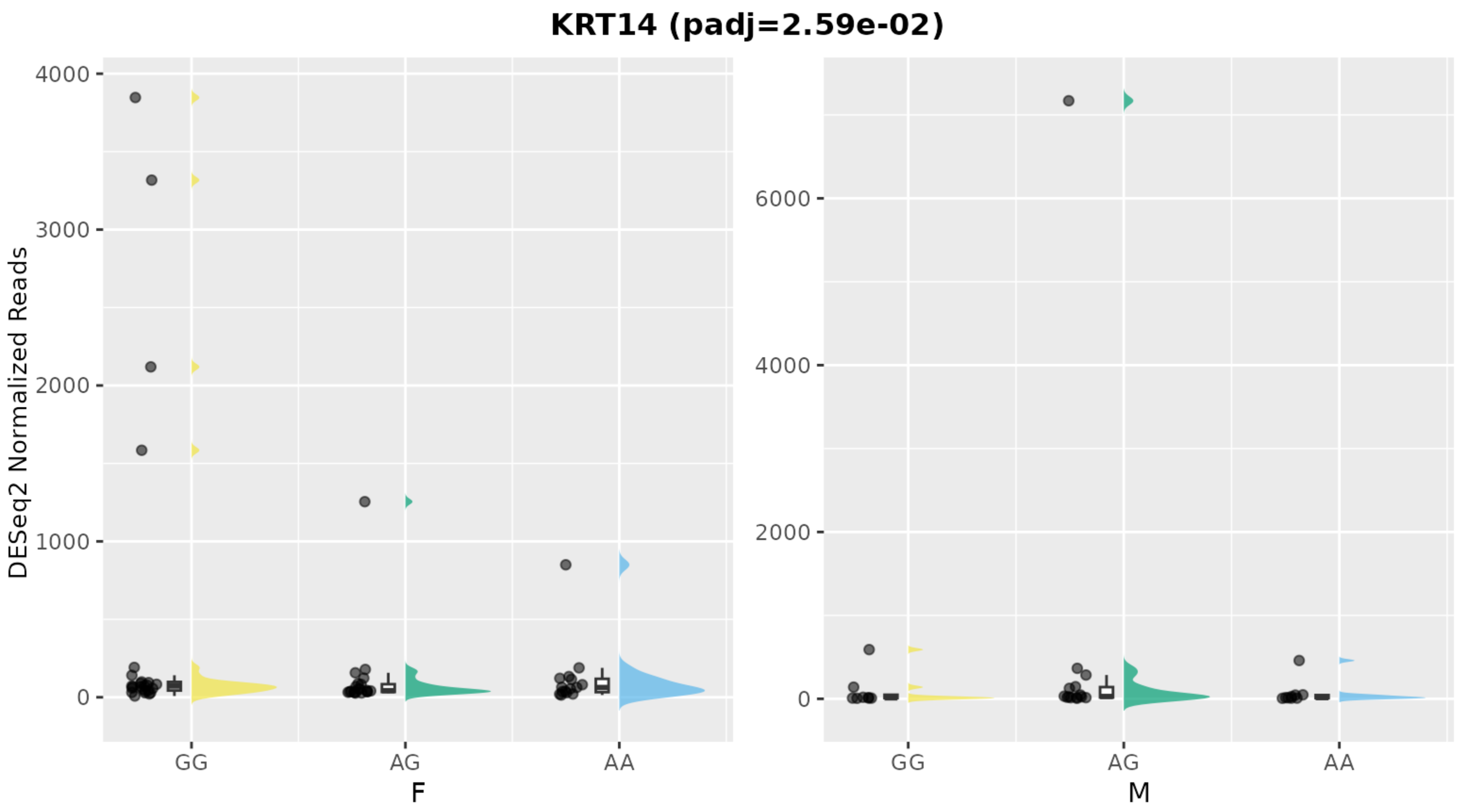
KRT14 (padj=2.59e-02)
DESeq2 Normalized Reads
4000
3000
2000
1000
0
6000
4000
2000
0
GG
AG
AA
F
GG
AG
AA
M

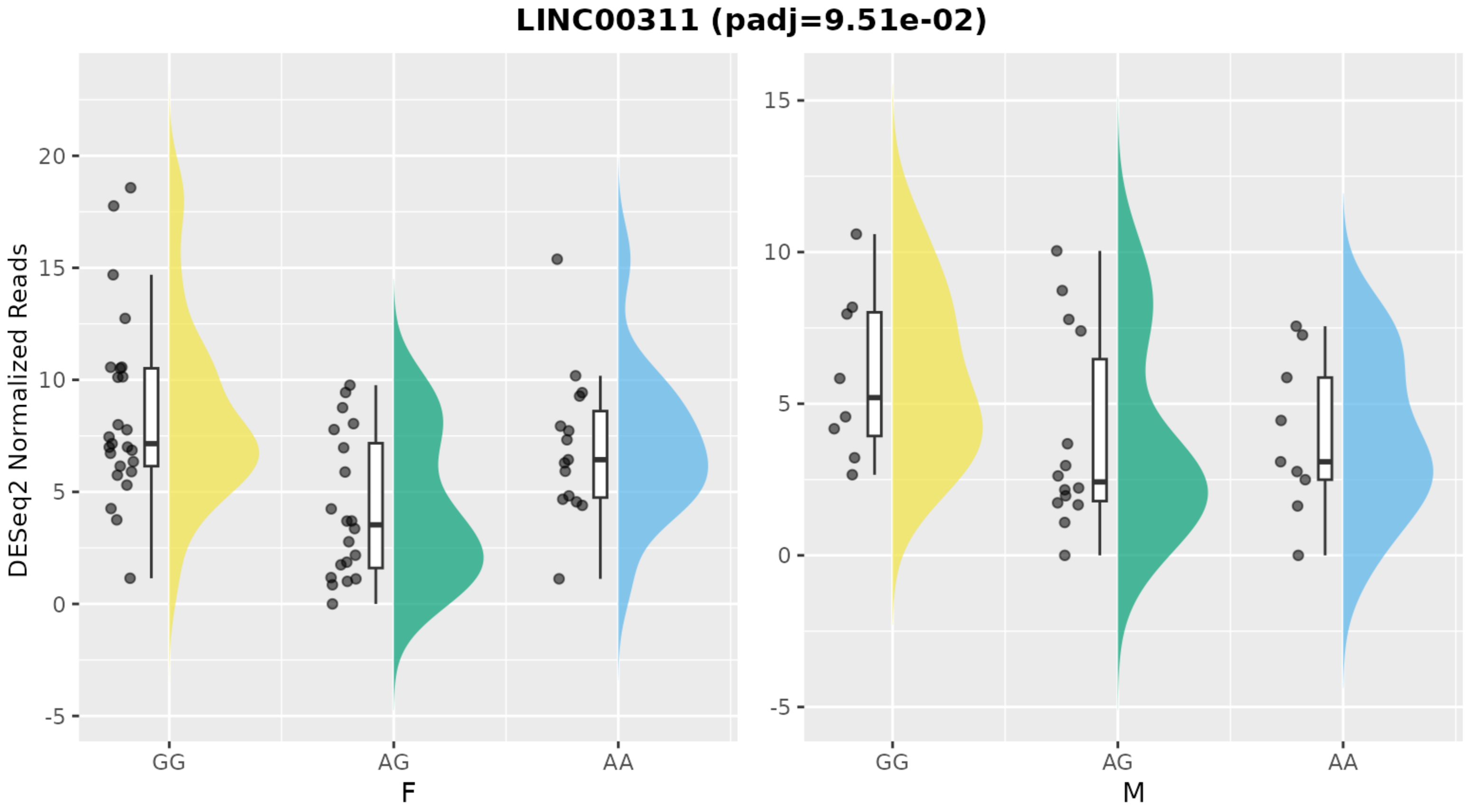

LINC00311 (padj=9.51e-02)
DESeq2 Normalized Reads
20
15
10
5
0
-5
GG
AG
AA
F
15
10
5
0
-5
GG
AG
AA
M

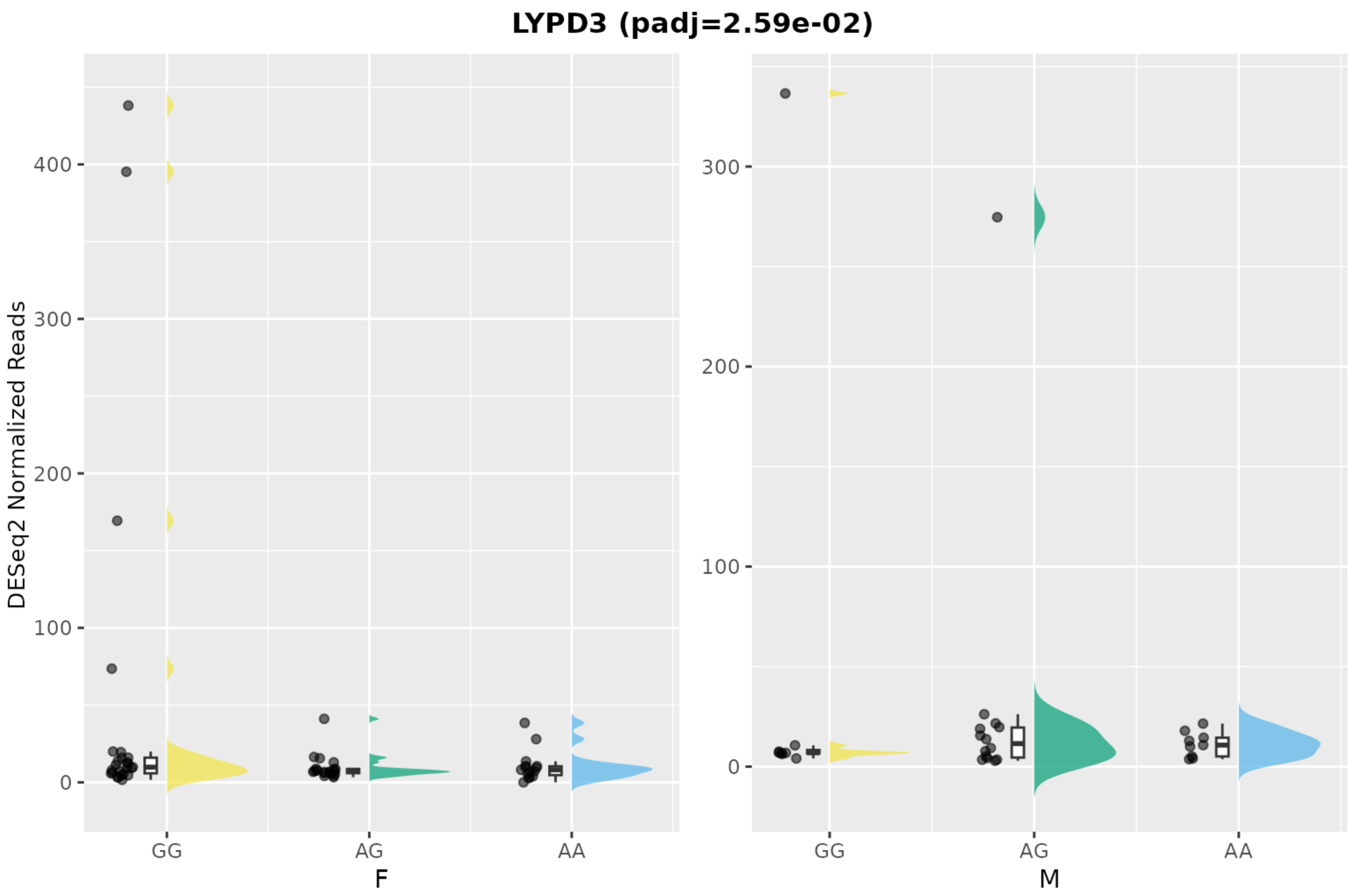

LYPD3 (padj=2.59e-02)
DESeq2 Normalized Reads
0
100
200
300
400
GG
AG
AA
F
0
100
200
300
GG
AG
AA
M

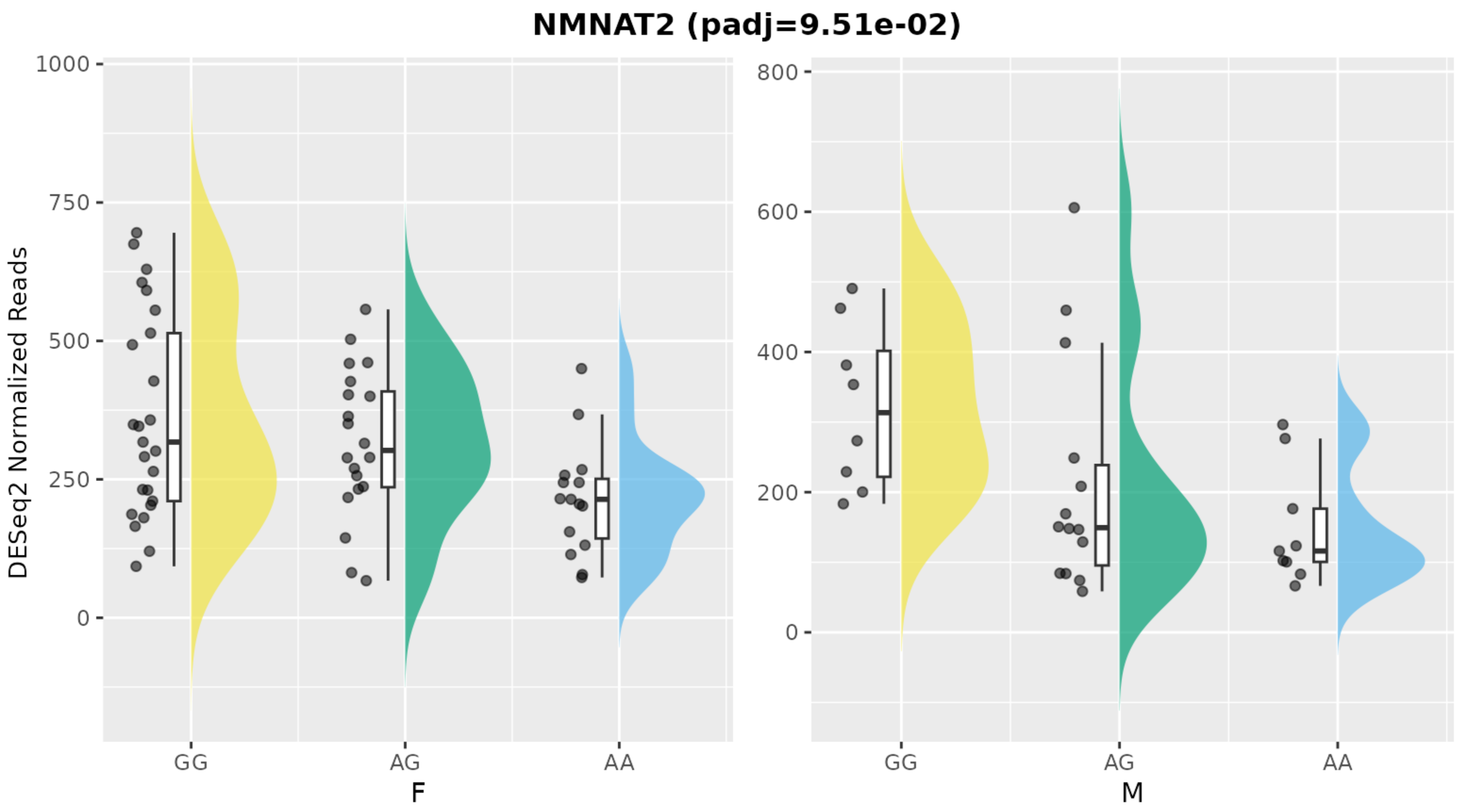
NMNAT2 (padj=9.51e-02)
DESeq2 Normalized Reads
1000
750
500
250
0
800
600
400
200
0
GG
AG
AA
F
GG
AG
AA
M

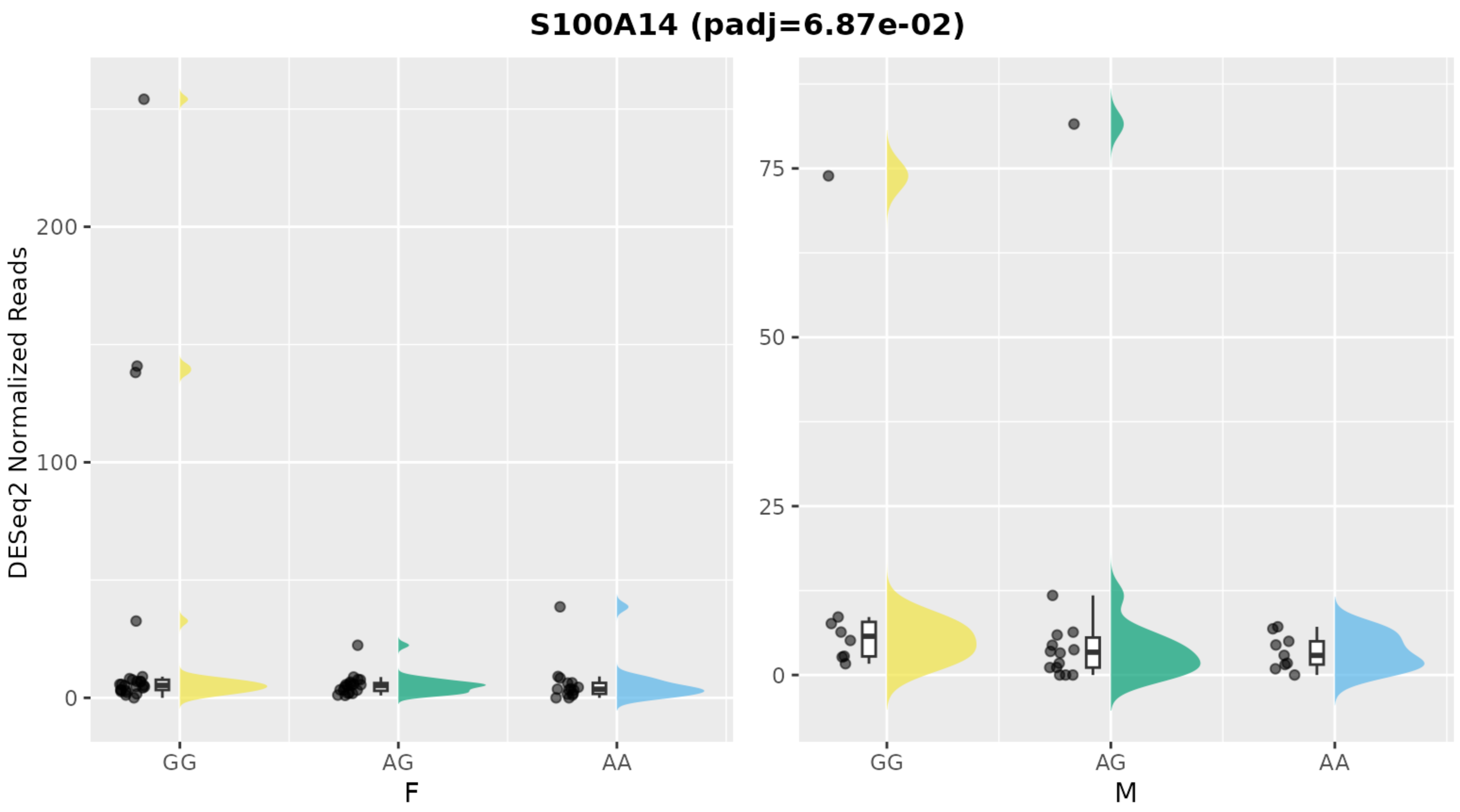
S100A14 (padj=6.87e-02)
DESeq2 Normalized Reads
200
100
0
GG
AG
AA
F
75
50
25
0
GG
AG
AA
M

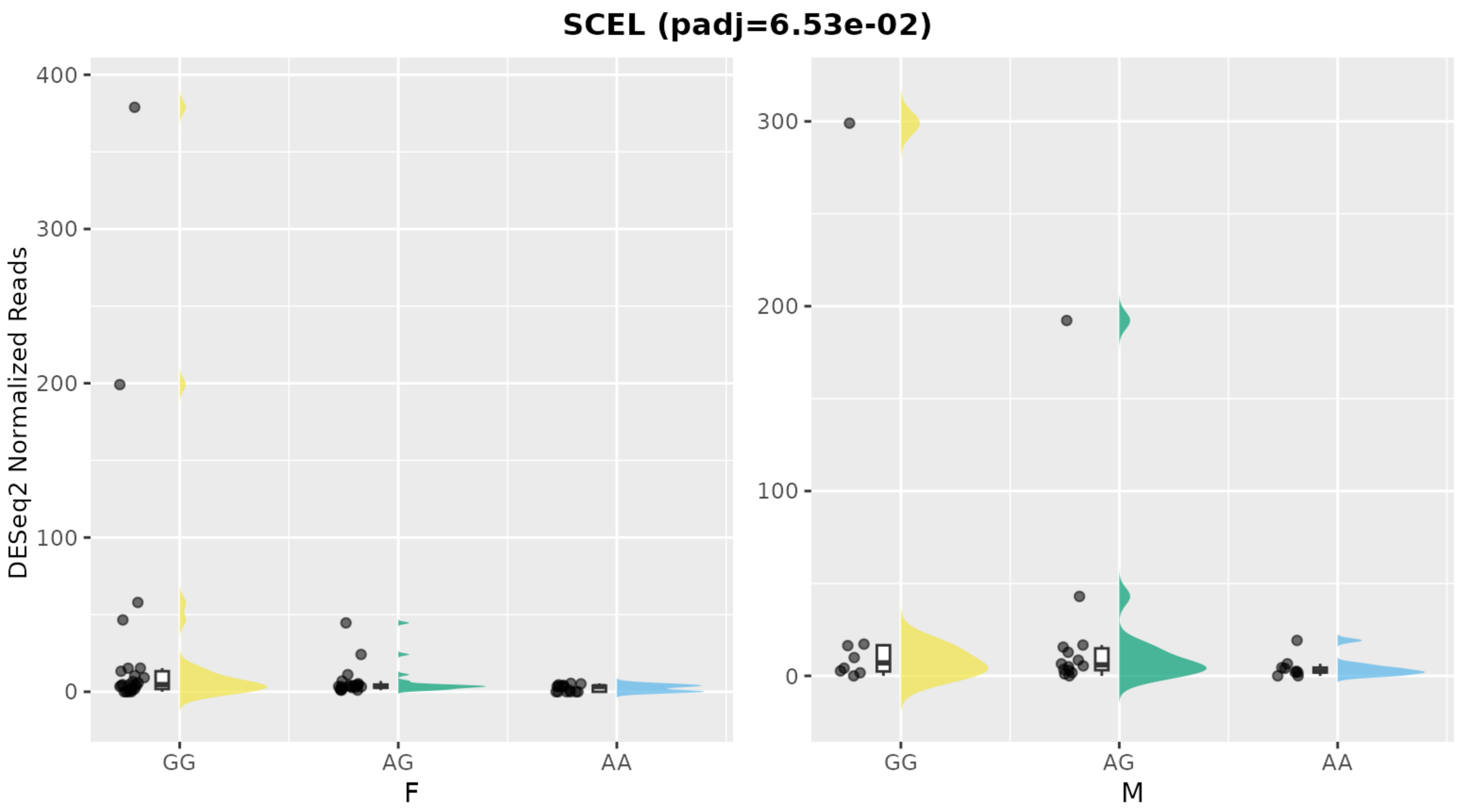
SCEL (padj=6.53e-02)
DESeq2 Normalized Reads
400
300
200
100
0
GG
AG
AA
F
M

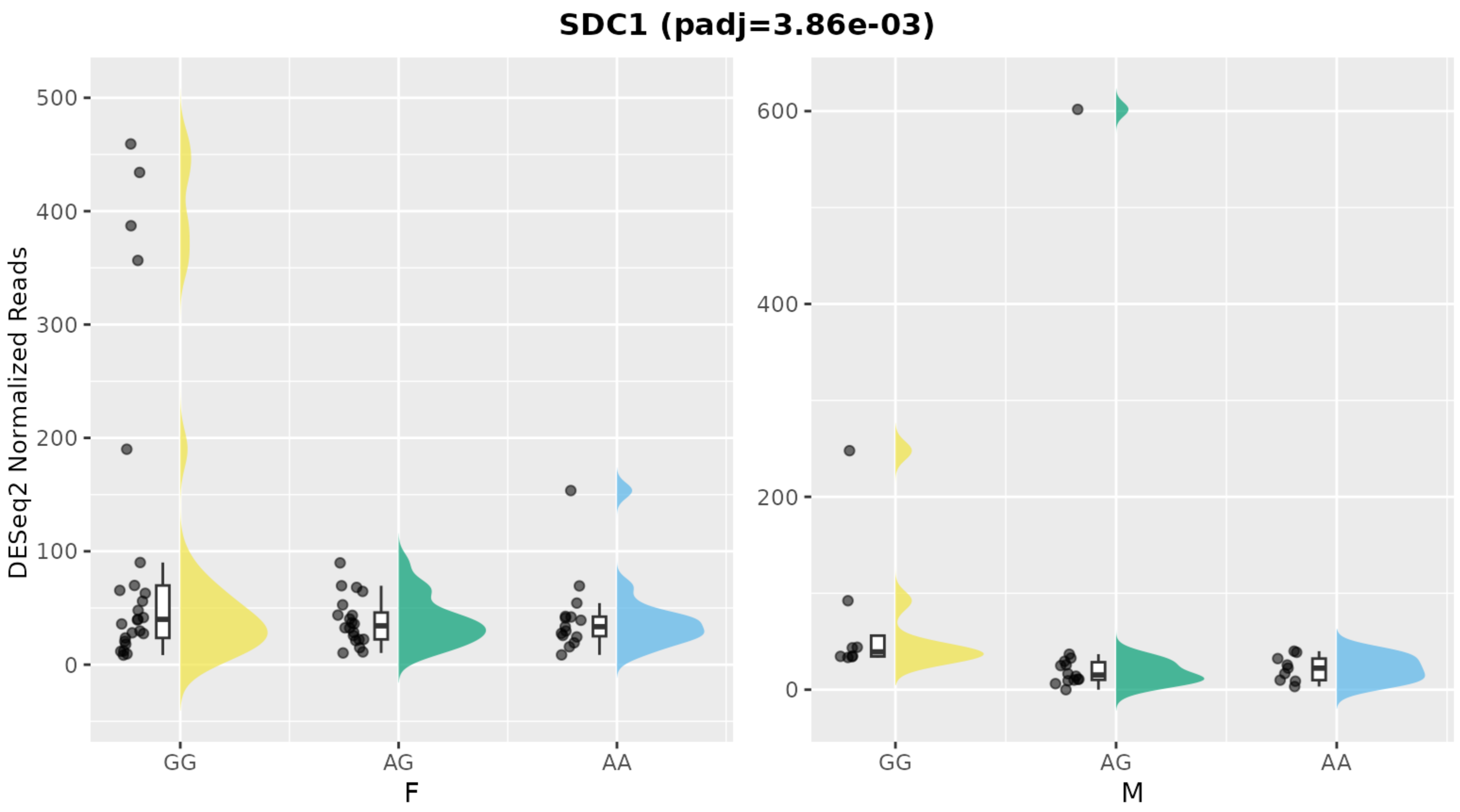

SDC1 (padj=3.86e-03)
DESeq2 Normalized Reads
500
400
300
200
100
0
600
400
200
0
GG
AG
AA
F
GG
AG
AA
M

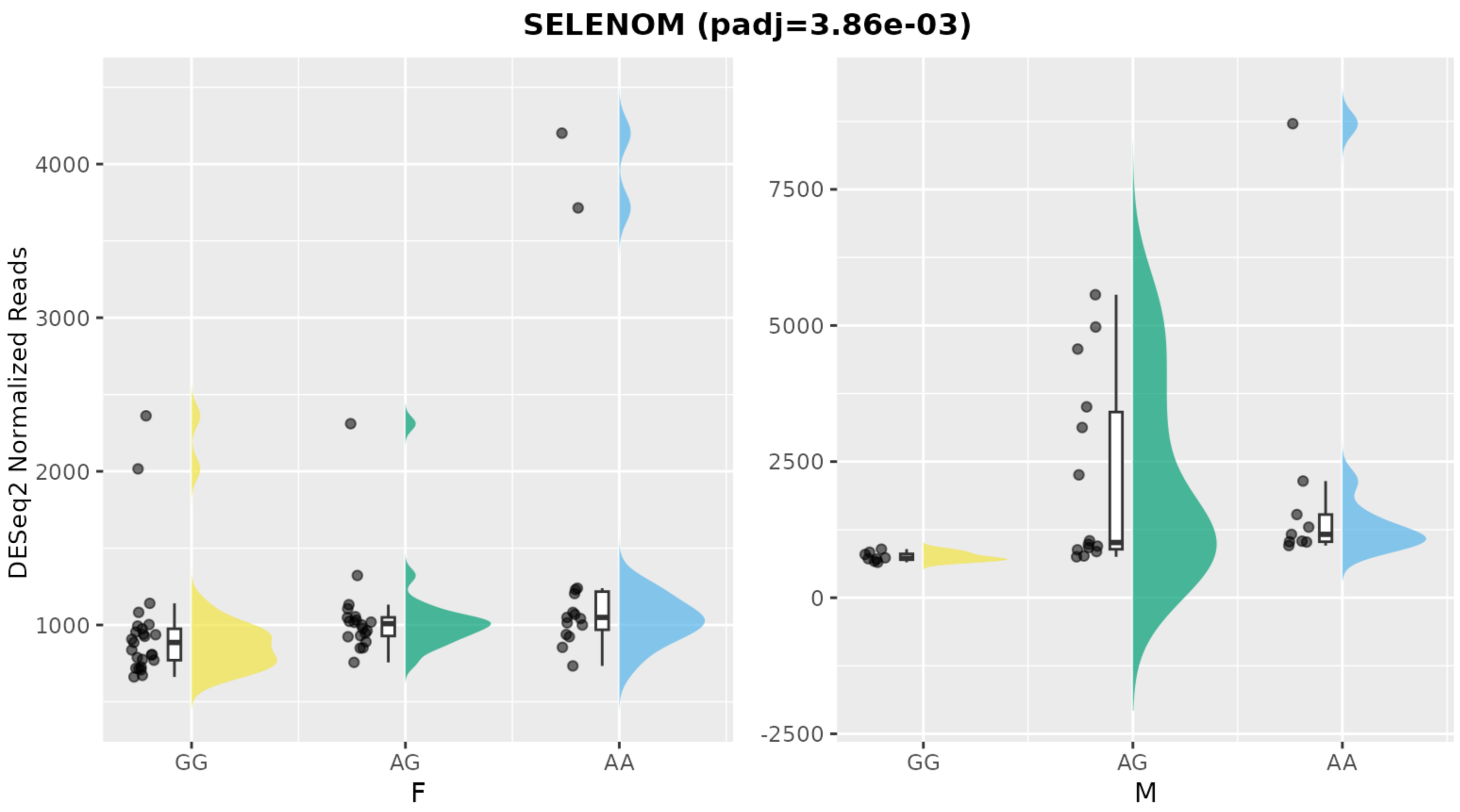
SELENOM (padj=3.86e-03)
DESeq2 Normalized Reads
4000
3000
2000
1000
GG
AG
AA
F
7500
5000
2500
0
-2500
GG
AG
AA
M

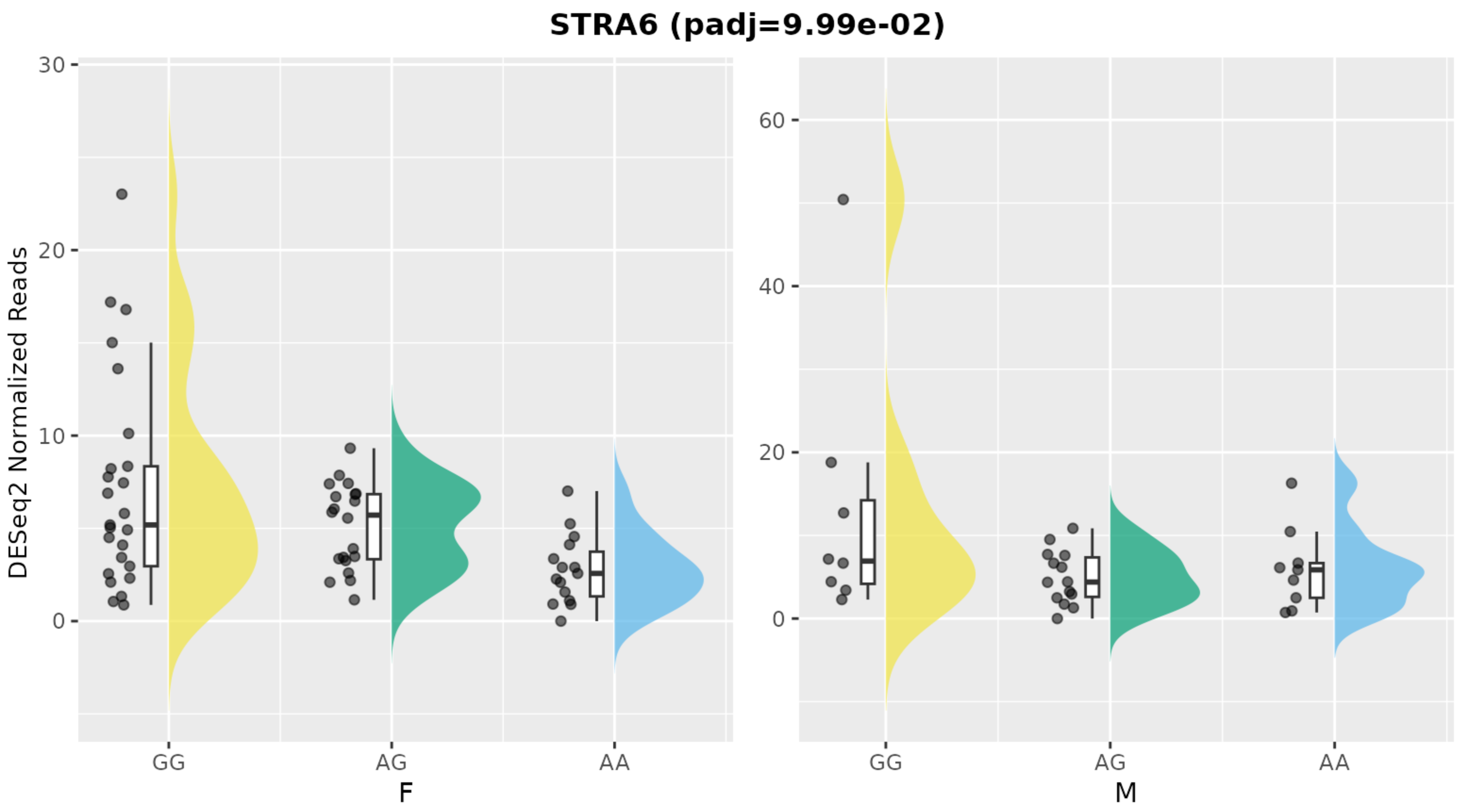
STRA6 (padj=9.99e-02)
DESeq2 Normalized Reads
30
20
10
0
60
40
20
0
GG
AG
AA
F
GG
AG
AA
M

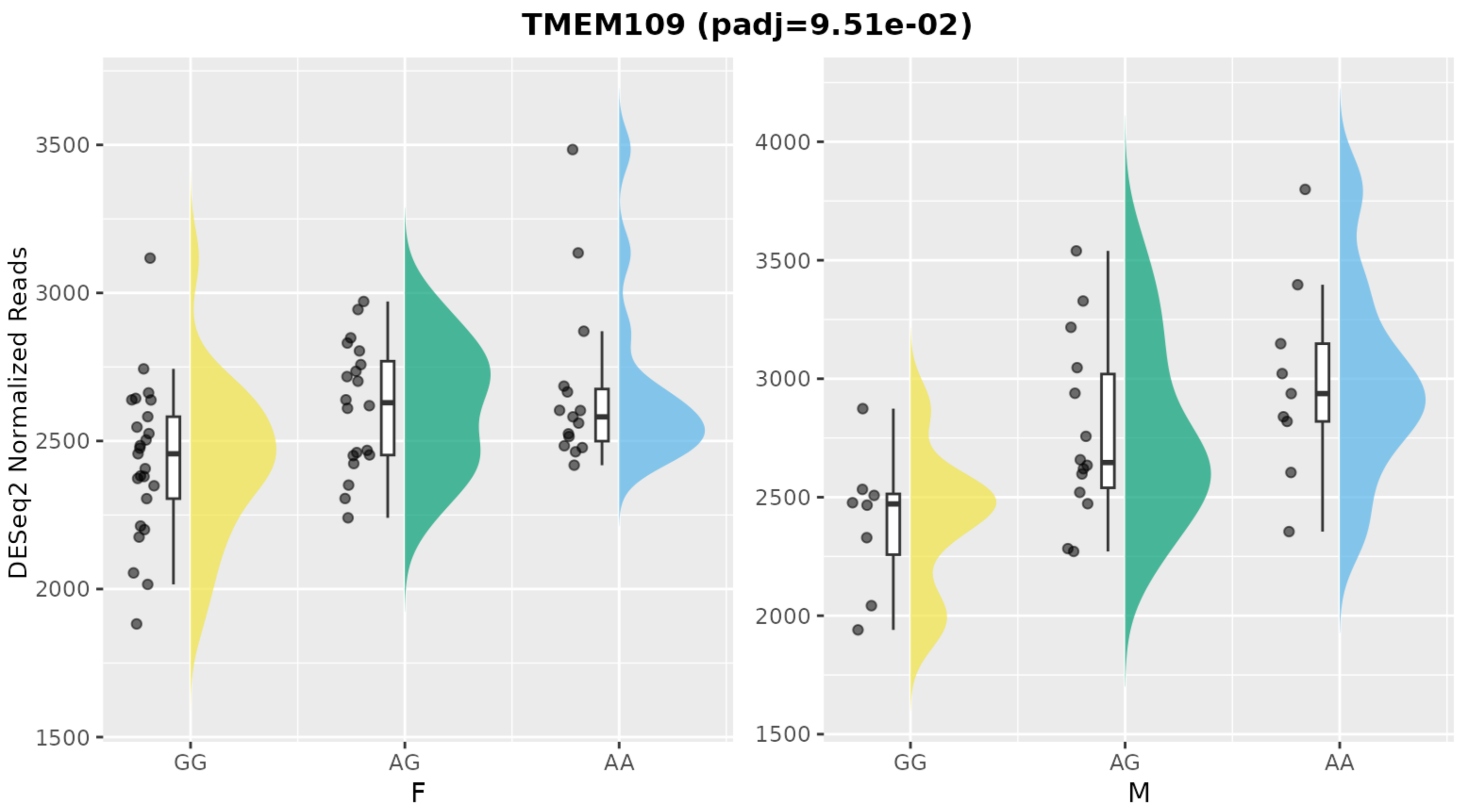


**Figure 3. Rain cloud plots of expression of the remaining 22 differentially expressed genes, stratified by sex and rs373863828 genotype.** Sex is indicated as F for female and M for male along the x-axis. Expression as measured by DESeq2 normalized reads. Quartiles and median of DESeq2 normalized reads for respective combination of rs373863828 genotype and sex are represented by the box plots, with individual samples represented by jittered points. Half-violin plots illustrate the shape of the distribution. Rain cloud plots for top two DEGs, *KRT1* and *KRT10,* are shown in **Figure 3**.
